# Revascularisation versus amputation for chronic limb-threatening ischaemia: a systematic review and meta-analysis of clinical outcomes and patient characteristics

**DOI:** 10.64898/2026.08.26.26361311

**Authors:** Jordan Luke Green, Henry Davies, David Alexander Russell

**Author notes:** **Corresponding Author & Address:** Jordan Green, Leeds Vascular Institute, Leeds Teaching Hospitals NHS Trust Leeds, LS1 3EX, United Kingdom.

## Abstract

**Background:** The relative merits of infrainguinal bypass and primary major lower limb amputation (MLLA) for chronic limb-threatening ischaemia (CLTI) remain uncertain, and the baseline profiles of patients selected for each strategy are poorly described.

**Methods:** A systematic review and meta-analysis were undertaken in accordance with PRISMA 2020 and prospectively registered (PROSPERO: CRD42022356094). MEDLINE, Embase, CENTRAL, and CINAHL were searched from inception to March 2025. Prospective studies of adults with CLTI undergoing primary infrainguinal bypass or primary MLLA were eligible. Mortality, major adverse cardiovascular events (MACE) and subsequent amputation outcomes were synthesised using random-effects meta-analysis of proportions. Baseline comorbidity profiles were also extracted.

**Results:** Twenty-seven studies involving 6,576 patients were included: 5,779 underwent infrainguinal bypass and 797 underwent MLLA. After bypass, pooled mortality was 3.7% at 30 days (95% CI 2.8%–4.9%, I² = 49.4%), 18.5% at 1 year (95% CI 15.6%–21.9%, I² = 62.3%), and 54.3% at 5 years (95% CI 50.5%–58.0%, I² = 0%). After MLLA, pooled mortality was 9.2% at 30 days (95% CI 4.1%–19.3%, I² = 73.5%), 28.5% at 1 year (95% CI 13.3%–51.0, I² = 70.8%), and 39.9% at 2 years (95% CI 0.3%–99.3, I² = 90.5%), although longer-term estimates were limited by sparse data and marked heterogeneity. Thirty-day MACE was 6.5% (95% CI 4.3%–9.7, I² = 63.5%) after bypass and 2.8% after MLLA (95% CI 0.1%–37.6%, I² = 0%). Early subsequent major amputation after bypass occurred in 3.9% of patients (95% CI 2.0%–7.7%, I² = 91.2%), rising to 16.2% at 1 year (95% CI 12.6%–20.5%, I² = 82.0%) and 33.3% at 3 years (95% CI 20.1%–49.8%, I² = 0%). Early re-amputation after MLLA occurred in 10.9% of patients (95% CI 4.5%–24.4%, I² = 40.3%). Baseline comorbidity burden was high in both groups, with substantial heterogeneity across studies.

**Conclusions:** CLTI carries a poor prognosis regardless of treatment strategy. Infrainguinal bypass is associated with lower early mortality and better early limb preservation than primary MLLA, but long-term survival remains poor and later limb failure is common. Primary MLLA is not a low-risk alternative. Better contemporary comparative evidence utilising modern causal inference approaches is needed to support individualised decision-making.

## Introduction

Chronic limb-threatening ischaemia (CLTI) represents the most severe manifestation of peripheral artery disease (PAD) and is associated with substantial morbidity, mortality, and healthcare burden. Characterised by ischaemic rest pain, gangrene, or ulceration attributable to peripheral arterial occlusive disease, CLTI threatens both limb viability and patient survival.^1^ Despite advances in revascularisation strategies, outcomes remain poor, with an estimated one quarter of patients dying and a further substantial proportion undergoing major lower limb amputation (MLLA) within one year of diagnosis.^2^

The primary therapeutic objective in CLTI is restoration of sufficient blood flow to achieve pain relief and tissue healing while preserving functional mobility. Contemporary management includes endovascular intervention, open surgical revascularisation, major amputation, or palliation, with treatment selection influenced by patient risk, anatomical complexity, limb severity, and expected functional outcomes.^1^ Although an endovascular-first strategy is increasingly adopted internationally for select groups of patients, open infrainguinal bypass remains an important treatment option for patients with complex femoropopliteal or infrapopliteal disease, particularly when endovascular intervention is unlikely to provide durable limb salvage.^3–5^

Major lower limb amputation remains an essential treatment for selected patients with CLTI where revascularisation is unlikely to provide meaningful benefit, including those with extensive tissue loss, non-functional limbs, prohibitive operative risk, or limited life expectancy.^1^ However, MLLA is associated with profound consequences for patients, including reduced mobility, impaired quality of life, and substantial mortality. Previous studies have reported mortality exceeding 30% within one year following amputation for CLTI, increasing with longer-term follow-up.^6^ Furthermore, although amputation may represent a definitive procedure for controlling infection or ischaemic pain, it is frequently perceived by patients as a devastating outcome, with many reporting fear of amputation comparable to or exceeding fear of death.^7^

The decision between attempting limb salvage with infrainguinal bypass or proceeding directly to MLLA is therefore complex. Current guidelines recommend consideration of patient-specific factors, including comorbidity burden, functional status, anatomical feasibility, and anticipated survival, when selecting treatment strategies.^1^ Several risk prediction models, including BASIL-Weibull, PREVENT-III, and FINNVASC, have been developed to estimate survival or amputation-free survival following revascularisation; however, their performance is generally modest, limiting their ability to guide individual treatment decisions reliably.^8–10^ Similarly, prediction models for perioperative mortality following MLLA demonstrate only moderate discrimination, highlighting the ongoing uncertainty in accurately identifying patients most likely to benefit from limb salvage versus primary amputation.^11^

Direct comparative evidence between infrainguinal bypass and MLLA is limited. Randomised trials comparing these strategies are unlikely to be feasible due to ethical concerns surrounding allocation to primary amputation when limb salvage may be achievable. Consequently, available evidence is derived predominantly from observational studies, which are vulnerable to confounding by indication. Patients selected for amputation are often thought to be older, frailer, and have greater comorbidity burden than those considered suitable for bypass; therefore, differences in outcomes may reflect underlying patient characteristics rather than the treatment strategy itself.

Despite these limitations, several large observational propensity-matched studies have suggested poorer survival following primary MLLA compared with infrainguinal revascularisation. Barshes et al. reported lower 30-day mortality following primary revascularisation compared with amputation in propensity-matched patients with CLTI (6.5% vs 10.0%).^12^ Similarly, Mustapha et al. demonstrated improved longer-term survival and reduced costs among patients undergoing revascularisation compared with MLLA within a large Medicare cohort.^13^ However, residual confounding remains an important limitation, as clinical assessments influencing treatment choice, including frailty, functional status, and perceived operative risk, are difficult to capture using routinely collected data.

Systematic reviews have previously compared endovascular intervention and open bypass for CLTI, demonstrating similar mortality and limb salvage outcomes but greater durability following surgical bypass.^14,15^ Separately, systematic reviews have quantified mortality following MLLA for all non-traumatic causes, demonstrating consistently poor long-term survival.^6^ However, no systematic review has directly compared outcomes following infrainguinal bypass versus MLLA in patients with CLTI, nor has synthesised differences in baseline patient characteristics between these treatment pathways. Understanding whether observed outcome differences are attributable to treatment choice or underlying differences in patient selection is essential to inform shared decision-making and optimise treatment allocation.

This systematic review therefore aimed to synthesise clinical outcomes following infrainguinal surgical bypass and MLLA in patients with CLTI and to examine differences in baseline characteristics between these treatment groups that may contribute to observed outcome variation.

## Methods

### Protocol Registration and Reporting Standards

This systematic review and meta-analysis was conducted according to the Preferred Reporting Items for Systematic Reviews and Meta-Analyses (PRISMA) 2020 statement. The review protocol was prospectively registered with the International Prospective Register of Systematic Reviews (PROSPERO; CRD42022356094).

### Eligibility Criteria

Studies were eligible for inclusion if they enrolled adults aged ≥18 years with chronic limb-threatening ischaemia (CLTI), defined as Rutherford category 4–6 or Fontaine stage III–IV disease. Eligible interventions included patients undergoing infrainguinal open surgical revascularisation involving the superficial femoral, popliteal, tibial, or pedal arteries. Comparator studies included primary major lower limb amputation (MLLA), defined as amputation above the ankle performed either as the initial treatment strategy or following failed endovascular intervention.

Randomised controlled trials, quasi-randomised trials, prospective cohort studies, prospective registries, and prospective observational studies were eligible for inclusion. Retrospective studies, review articles, case reports, case series, and studies with fewer than 50 patients in either treatment group were excluded. Studies involving isolated aortoiliac, common femoral, or profunda femoris artery revascularisation were excluded, as were studies of intermittent claudication unless outcomes for patients with CLTI were reported separately. Only studies published in English were included.

### Information Sources and Search Strategy

A comprehensive literature search was undertaken in MEDLINE (1946 onwards), Embase (1947 onwards), the Cochrane Central Register of Controlled Trials (CENTRAL), and CINAHL from database inception to March 2025. MEDLINE, Embase, and CENTRAL were searched through the Ovid interface, whereas CINAHL was searched through EBSCOhost.

Search strategies combined controlled vocabulary terms and free-text keywords relating to CLTI, critical limb ischaemia, peripheral arterial disease, infrainguinal bypass surgery, arterial revascularisation, and MLLA (Supplementary Material 1). Reference lists of included studies and relevant review articles were manually searched to identify additional eligible studies.

### Study Selection

Search results were imported into Rayyan (Rayyan Systems Inc., Cambridge, MA, USA) for screening and duplicate removal. Study selection was performed in two stages. First, two reviewers independently screened titles and abstracts against the predefined eligibility criteria. Potentially relevant articles then underwent full-text review by the same reviewers. Disagreements at either stage were resolved through discussion and consensus with an additional third reviewer. Reasons for exclusion at the full-text stage were recorded.

### Data Extraction

Data extraction was performed independently using a pre-specified and piloted data extraction form. Discrepancies were resolved by discussion and consensus.

Extracted study-level data included author, publication year, country, study design, recruitment setting, sample size, and duration of follow-up. Participant characteristics included age, sex, CLTI severity, comorbidities, and anatomical distribution of arterial disease. Treatment-related variables included type of revascularisation or amputation and relevant procedural details, where reported.

Outcome data were extracted at 30 days, 1 year, and annually thereafter up to 5 years when available. Extracted outcomes included all-cause mortality, major adverse cardiovascular events (MACE) and amputation or re-amputation. When multiple publications reported overlapping cohorts, the study providing the most complete dataset was included; authors were contacted when overlap could not be determined with certainty. In addition, baseline patient characteristics and comorbidity profiles were collected to explore differences between populations undergoing primary revascularisation and primary amputation.

### Outcomes

The primary outcome was all-cause mortality at 30 days and annually up to 5 years following intervention. Secondary outcomes included: MACE, defined as non-fatal cardiac event or stroke; amputation (in bypass cohorts), defined as a major amputation above the ankle; and re-amputation (for amputation cohorts), defined as ipsilateral revision of the primary amputation to a higher level.

### Risk of Bias Assessment

Randomised controlled trials were evaluated using the revised Cochrane Risk of Bias tool for randomised trials (RoB-2).^16^ Prospective non-randomised studies, registries, and cohort studies were assessed using the Newcastle-Ottawa Scale (NOS).^17^

### Data Synthesis and Statistical Analysis

A narrative synthesis was undertaken to describe differences in study settings, patient populations, disease severity, and treatment strategies between cohorts undergoing primary revascularisation and MLLA.

Where sufficient clinical and methodological homogeneity existed, quantitative synthesis was performed. Where outcome data were reported as percentages only, event counts were derived from the reported percentage and study denominator. Pooled estimates of mortality and baseline comorbidities were calculated separately for revascularisation and amputation cohorts using random-effects meta-analysis of proportions. Proportions were pooled using the DerSimonian–Laird random-effects model and presented with corresponding 95% confidence intervals. Cumulative incidence estimates were reported for non-comparative studies.

Statistical heterogeneity was assessed using the I² statistic, with values interpreted according to established guidance. Given the anticipated differences in study design, patient selection, and outcome reporting, random-effects models were selected a priori. No subgroup analyses were planned.

All statistical analyses were performed using R version 2026.07.0+139 (R Foundation for Statistical Computing, Vienna, Austria). Meta-analyses were conducted using the meta and metafor packages.

## Results

### Study Selection (Figure 1)

7898 articles were screened according to the strict inclusion and exclusion criteria, resulting in the selection of 30 articles, encompassing 27 individual studies. Three studies were reported across two articles each, with a further one article describing two treatment cohorts within the one study, and therefore a total of 28 cohorts were included.

**Figure 1.**
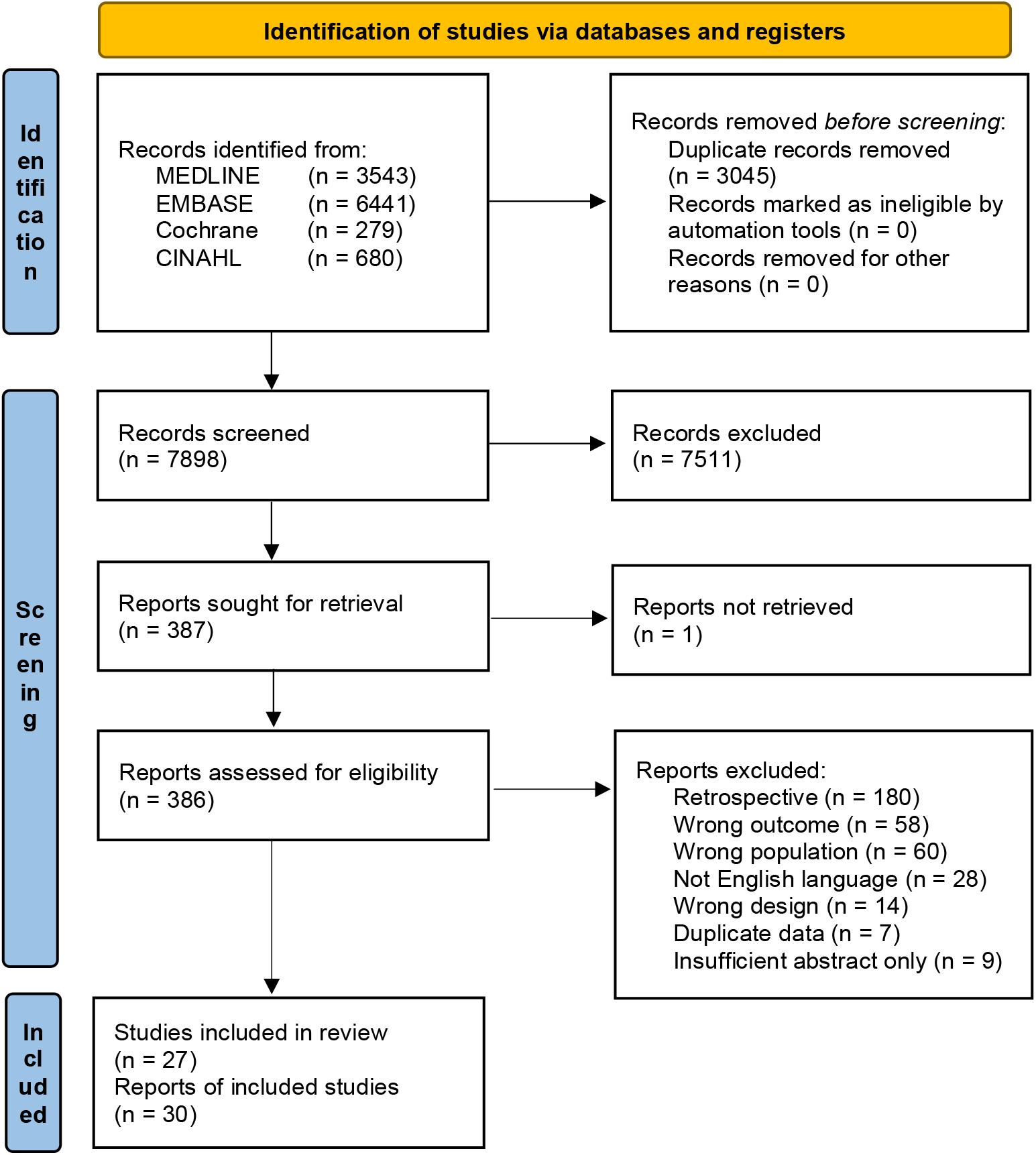
PRISMA flow diagram for identification and inclusion of studies for the systematic review.

Seven study records were excluded that met inclusion criteria as they were duplicate datasets. Preliminary results for the LIMBSAVE registry from Troisi et al., a registry of in-situ infrainguinal great saphenous vein bypass for CLTI^18^ were superseded by the 2-year results.^19^ Similarly, the interim analysis of the CRITISCH registry of the management from Bisdas et al.^20^ was superseded by the one year results by Stavroukalis et al.^21^ Soderstrom et al. reported overlapping patients in two papers assessing healing time of ulcers after infrainguinal bypass for CLTI;^22,23^ the earlier paper was a larger dataset that included data on outcomes for the systematic review, with no additional value from the 2009 paper which was therefore excluded. Multiple papers on the BEST-CLI trial were also available; Farber et al. and Conte et al. provided the scope of outcome data needed for this review,^5,24^ but the additional analyses presented by Giles et al.,^25^ Farber et al.^26^ and Ulloa et al.^27^ did not present any further data and were therefore excluded. Finally, the publication of results from the BASIL-2 trial were comprehensive from the original Bradbury et al. publication,^4^ with no additional benefit for this review from Moakes et al., thus resulting in paper exclusion.^28^

### Study Characteristics (Table 1)

A total of 27 studies met the inclusion criteria, comprising 21 infrainguinal bypass cohorts and seven MLLA cohorts. Across all included studies, 6,576 patients were analysed, of which 5,779 (87.9%) underwent infrainguinal bypass and 797 (12.1%) underwent MLLA. Studies were published between 1984 and 2025 and originated from Europe, North America, South America, Africa and Asia. The evidence base consisted predominantly of prospective observational studies, with nine randomised controlled trials evaluating infrainguinal bypass. Follow-up ranged from 30 days to 6 years.

**Table 1.** Summary of all included studies within the systematic review.

| Study | Study Type | Setting | Recruitment | Number of Patients | Indication for Intervention | Follow-Up |
| --- | --- | --- | --- | --- | --- | --- |
| *Infrainguinal Bypass* |  |  |  |  |  |  |
| Attia 2014 | Prospective registry of consecutive patients | Single-centre, Egypt | Jan 2010 to Dec 2012 | 50 | Claudication (10%)<br>Rest pain (18%)<br>Tissue loss (72%) | 6 weeks, then 3, 6, 9, 12 and 18 months |
| BASIL Trial Participants 2005, Bradbury 2010 | Randomised controlled trial | 27 centres, UK | Aug 1999 to Jun 2004 | 195 | Rest pain (27%)<br>Tissue loss (73%) | Up to 3-years |
| Bradbury 2023 | Randomised controlled trial | 41 centres, Europe | Jul 2014 to Nov 2020 | 172 | Rest pain only (13%)<br>Tissue loss only (23%)<br>Both rest pain and tissue loss (64%) | 1, 6, 12 and 24-months then annually (minimum of 2 years, median 40 months) |
| Darke 1989 | Randomised controlled trial | 7 centres, UK | Not stated | 59 | Tissue loss (44%)<br>Rest pain (56%) | 12 months |
| Dick 2007 | Prospective cohort | Single centre, Switzerland | Jan 1999 to Jun 2004 | 85 | Rutherford 4 (20.0%)<br>Rutherford 5 (76.5%)<br>Rutherford 6 (3.5%) | 2, 6 and 12 months |
| Farber 2022, Conte 2025 | Randomised controlled trial | 150 centres, International | Aug 2014 to Oct 2019 | 857 | CLTI - not described further | Median 2.7 years |
| Gloviczki 1994 | Prospective cohort | Single-centre, USA | Sep 1987 to Jul 1993 | 100 | Rest pain (9%)<br>Tissue loss (91%) | Mean 2.1 years |
| Jensen 2007 | Randomised controlled trial | 13 centres across Denmark, Norway and Finland | Oct 1993 to Jan 1997 | 119 | CLTI - not described further | 24 months |
| Kapfer 2006 | Randomised controlled trial | 19 centres, Germany | Jun 1995 to Nov 1998 | 265 | Claudication (6.8%)<br>Rest pain (32.8%)<br>Tissue loss (60.4%) | 36 months |
| Lundgren 2013 | Randomised controlled trial | 29 centres, Sweden | Not stated | 329 | Rest pain or tissue loss (94%) | 1, 3, 6, and 12 months then annually to max of 5 years |
| Mezzetto 2024 | Prospective registry of consecutive patients | Multi-centre, Italy | From 2020 | 68 | Rutherford 3 (8.8%)<br>Rutherford 4 (35.3%)<br>Rutherford 5 (29.7%)<br>Rutherford 6 (16.2%) | 1, 6, 12-months then annually |
| Nguyen 2006, Conte 2006 | Randomised controlled trial | 83 centres, USA | Nov 2001 to Oct 2003 | 1404 | Rest pain (25%)<br>Tissue loss (75%) | 12 months |
| Panayiotopoulos 1997 | Prospective cohort | Single centre, UK | Jun 1991 to Dec 1994 | 109 | Rest pain (34.9%)<br>Tissue loss (65.1%) | 3, 6, 12-months then 6-monthly after (up to 42-months) |
| Robbs 1984 | Prospective cohort | Single-centre, Durban, South Africa | Not described | 113 | Pre-gangrene (20%)<br>Gangrene (80%) | 30 days then up to 3-years |
| Sarac 1998 | Randomised controlled trial | 2 centres, Florida, USA | Jan 1993 to Sep 1995 | 56 | Rest pain or tissue loss (95.3%) | Up to 42 months |
| Soderstrom 2008 | Prospective cohort | Single-centre, Finland | Jan 2005 to Jul 2006 | 184 | Fontaine IV (100%) | 1-year |
| Stavroulakis 2018 | Prospective registry of consecutive patients | 27 centres, Germany | Jan 2013 to Sep 2014 | 284 | Rutherford 4 (25%)<br>Rutherford 5 (49%)<br>Rutherford 6 (25%) | 1-year |
| Takeji 2018 | Prospective cohort | Single-centre, Japan | Jan 2010 to Jan 2016 | 157 | Rest pain (24%)<br>Tissue loss (76%) | Up to 2-years |
| Troisi 2022 | Prospective registry of consecutive patients | 43 centres, Italy | Jan 2018 to Dec 2019 | 541 | Tissue loss (67.5%)<br>Rest pain (32.5%) | Mean 12.1 months |
| Watson 1999 | Prospective cohort | 21 centres across Europe | Not described | 507 | Fontaine III (34%)<br>Fontaine IV (65%) | 14-days, 3-months and 12-months. |
| Wolfe 2000 | Prospective cohort | Single-centre, Germany | May 1986 to 1990 | 125 | Rest pain (2.3%)<br>Gangrene (97.7%) | 30-days then up to 6-years |
| *Amputation* |  |  |  |  |  |  |
| Cosgrove 2002 | Prospective cohort | Single-centre, UK | Aug 1992 to Jan 1996 | 217 | CLTI - not described further | 30 days |
| Eneroth 1992 | Prospective cohort | 5 centres, Sweden | Jan 1987 to Mar 1988 | 177 | Progressive gangrene, with or without septicaemia, or intractable pain (100%) | 2-years |
| Matielo 2008 | Prospective cohort | Single-centre, Brazil | Sept 2004 to Mar 2006 | 56 | Tissue loss (100%) | 30-days |
| Ploeg 2005 | Prospective cohort | Single-centre, Netherlands | Jan 1996 to Dec 2002 | 97 | CLTI - not described further | Up to 5-years |
| Robbs 1984 | Prospective cohort | Single-centre, South Africa | Not described | 88 | Established gangrene (94%) | Up to 3-years |
| Torbjoernsson 2022 | Prospective cohort | Single-centre, Sweden | Sept 2014 to May 2018 | 73 | CLTI - not described further | 12 months |
| Wolthius 2006 | Prospective cohort | Single-centre, UK | Jan 2001 to Dec 2003 | 89 | Tissue loss (100%) | 30 days |

#### Infrainguinal Bypass - Randomised Controlled Trials

Nine randomised controlled trials comprising 3,456 patients evaluated infrainguinal bypass. These studies included multicentre national and international collaborations and represented the highest-quality evidence identified within the review.

Early randomised evidence was provided by Sarac et al., who randomised 56 patients across two centres in Florida to either aspirin alone or warfarin and aspirin in high risk infrainguinal grafts between 1993 and 1995,^29^ and Darke et al., who enrolled 59 patients across seven UK centres randomising them to either popliteal or distal bypass targets.^30^ Both studies included patients presenting with rest pain and tissue loss and reported follow-up to at least 12 months.^29,30^ Larger European trials with longer follow-up subsequently expanded the evidence base: Kapfer et al. randomised 265 patients across 19 German centres to two different types of synthetic grafts for femoro-AT bypasses,^31^ while Jensen et al. recruited 119 patients from 13 centres across Denmark, Norway and Finland and assigned them to different synthetic grafts for above-knee femoro-popliteal bypass.^32^ In addition, Lundgren et al. contributed 329 patients randomised to different synthetic graft strategies for either femoro-popliteal or femoro-distal bypasses from 29 Swedish centres, and provided follow-up extending to 5 years.^33^

Four landmark contemporary randomised trials also contributed substantially to the pooled population. The BASIL trial enrolled 195 patients from 27 UK centres between 1999 and 2004 and followed participants for up to 3 years, randomising patients with all infrainguinal disease to either a bypass-first or endovascular-first strategy, with bypass patients receiving either vein or synthetic grafts.^3,34^ More recently, BASIL-2 recruited 172 patients from 41 European centres between 2014 and 2020, with a median follow-up of 40 months, examining patients with infrageniculate disease only and randomising them to a bypass-first or endovascular-first strategy.^4^

Further contemporary randomised evidence was provided by the PREVENT-III and BEST-CLI trials. BEST-CLI recruited 857 patients across 150 international centres between 2014 and 2019, with patients included in two cohorts (suitable autologous venous conduit available or not), with both cohorts randomised to either infrainguinal bypass or endovascular therapy.^5,24^ The PREVENT III study represents the largest prospective bypass population studied, comprising 1,404 patients across 83 centres in the United States, with patients randomised to either infrainguinal bypass using standard autologous vein conduit or edifoligide-administered vein conduit.^35,36^

#### Infrainguinal Bypass - Prospective Cohort and Registry Studies

Eight prospective cohort studies and four registry analyses contributed a further 2,323 bypass procedures. These studies were geographically diverse and included single-centre, national multicentre, and international collaborations.

Among the earliest studies, Robbs et al. reported a prospective cohort of 113 patients presenting to a single vascular service in South Africa and compared this to a cohort of MLLA. 80% of bypass patients had gangrene with 84% of bypasses to the popliteal artery and 16% to the tibial arteries, reflecting vascular practice at that time.^37^ Panayiotopoulos et al. also reported a consecutive series of 109 femoro-tibial bypasses performed at a single UK centre, with the aim to assess if bypass level was associated with subsequent knee salvage if amputation was required, with outcomes of amputation and survival. All patients had rest pain and 65.1% had tissue loss.^38^ In addition, Gloviczki et al. reported a cohort of 100 patients with CLTI treated with bypass to the pedal vessels in a single. As expected with the aggressive bypass target, 92% of patients had tissue loss.^39^

Soderstrom et al. reported a well-represented prospective cohort of 184 patients treated by infrainguinal bypass in a single Finnish vascular unit over 19 months. 46% of patients had gangrene and 68% of bypass grafts were to tibial or pedal vessels.^22^ Wolfle et al. described 125 German patients undergoing distal bypass with vein graft. The cohort were predominantly gangrenous disease (97.7%) and followed for up to 6 years.^40^

Dick et al. reported outcomes from a small cohort of 85 patients, 52% of which received a vein graft. 80% of the cohort had tissue loss, with follow-up to a year.^41^ Takeji et al. contributed a Japanese cohort of 157 patients who were compared to endovascular treatment, three-quarters of whom presented with tissue loss.^42^ Moreover, Watson et al. contributed one of the largest observational cohorts, comprising 507 patients from 21 European centre; all bypasses involved an above knee origin to a below-knee target bypass (82% vein, 18% prosthetic), and were followed up for a year.^43^ Furthermore, Mezzetto et al. reported outcomes from an Italian multicentre registry initiated in 2020, with 68 patients receiving a below knee bypass using a heparin-bonded synthetic graft,^44^ while Attia et al. described a prospective Egyptian registry of 50 patients undergoing infrainguinal bypass with any conduit.^45^

Larger registry-based investigations have also increasingly characterised contemporary bypass practice. Stavroulakis et al. reported the one-year outcomes of the CRITISCH registry, a multicentre prospective registry of patients with new CLTI across 27 German vascular centres over a 19-month period.^21^ 284 were treated with infrainguinal bypass. Major tissue loss was reported in 25%, 93% had complex occlusive disease (TASC C/D)^2^ and only 26% of bypasses were above the knee.^21^ Troisi et al. reported the 2-year outcomes of the LIMBSAVE registry, a prospective, multicentre registry of 541 patients with CLTI treated with in-situ vein infrainguinal bypass, using the LeMaitre^®^ valvulotome to destroy the vein valve cusps, across 43 Italian vascular centres. 67.5% of patients had tissue loss with 61.9% of bypasses to the popliteal artery and 38.1% to more distal vessels.^19^

#### Major Lower Limb Amputation - Randomised Controlled Trials

No randomised controlled trials evaluating primary major lower limb amputation met the eligibility criteria. Consequently, all evidence regarding amputation outcomes was derived from prospective observational studies.

#### Major Lower Limb Amputation - Prospective Cohort and Registry Studies

Seven prospective observational studies comprising 797 patients evaluated outcomes following MLLA. Compared with the bypass literature, these studies were generally smaller, predominantly single centre in design, and less likely to provide long-term follow-up.

The earliest evidence was provided by Robbs et al. in South Africa and Eneroth et al. in Sweden. Robbs et al. reported outcomes in 88 patients with established gangrene,^37^ while Eneroth et al. studied 177 patients with progressive gangrene, sepsis or intractable pain and provided follow-up extending to 2 years.^46^ Subsequent cohorts from the United Kingdom included Cosgrove et al., who reported 217 amputations between 1992 and 1996,^47^ and Wolthuis et al., who evaluated 89 patients with tissue loss between 2001 and 2003.^48^ Ploeg et al. contributed one of the few studies with longer-term follow-up, reporting outcomes from 97 Dutch patients followed for up to 5 years.^49^ Matielo et al. evaluated 56 patients with tissue loss,^50^ whereas Torbjörnsson et al. reported outcomes in 73 patients undergoing major amputation between 2014 and 2018.

Across all amputation studies, the underlying indication was almost universally advanced CLTI characterised by tissue loss, progressive gangrene, established gangrene or unsalvageable limb ischaemia. Disease severity reporting was generally less detailed than in bypass studies including the paucity of reporting disease location, and contemporary classification systems such as Rutherford or WIfI were rarely used.

### Risk of Bias

#### Randomised Controlled Trials of Infrainguinal Bypass (Figure 2)

Risk of bias among the randomised controlled trials was generally low to moderate. Two publications arising from the PREVENT III trial^35,36^ were judged to be at low risk of bias across all assessed domains. Similarly, BASIL was considered to be at low overall risk of bias for mortality outcomes; although treatment allocation was not blinded, analyses were performed according to the intention-to-treat principle and the principal outcomes relevant to this review, including mortality and major amputation, and were therefore objective clinical endpoints unlikely to be influenced by knowledge of treatment assignment.

**Figure 2.**
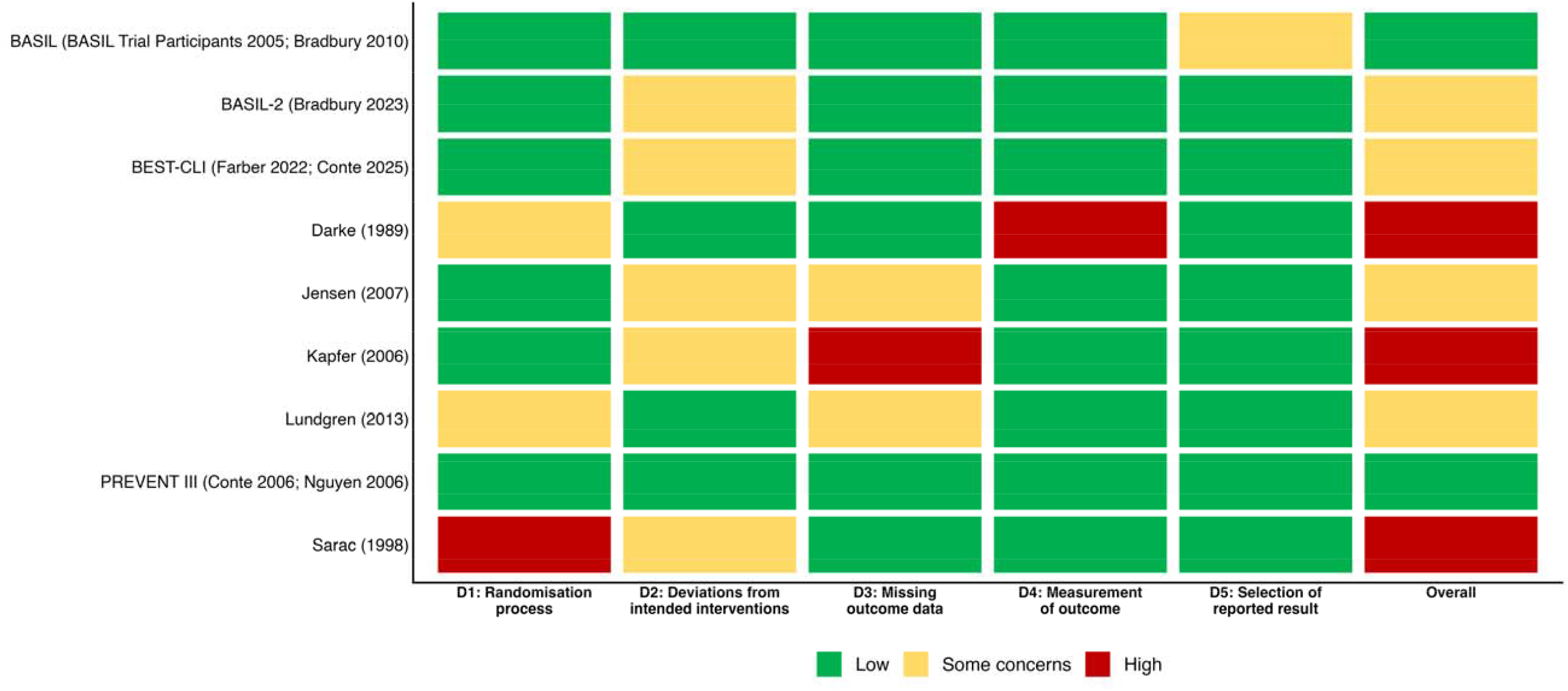
Risk of bias assessment using the Revised Risk of Bias tool (ROB-2) for randomised controlled trials of infrainguinal bypass.

Four studies were judged as having some concerns overall. Jensen et al. had concerns related primarily to deviations from intended interventions as well as limited reporting regarding missing outcome data.^32^ Lundgren et al. was judged to have some concerns relating to both the randomisation process and incomplete reporting of missing data. As with several studies included in the review, treatment allocation was performed using sealed envelopes, a method susceptible to subversion if allocation concealment is inadequately maintained, potentially allowing investigators to predict or influence treatment assignment.^33^ BASIL-2 and BEST-CLI were also judged as having some concerns overall. In both trials, the principal source of potential bias arose from deviations from intended interventions. These pragmatic, open-label revascularisation strategy trials necessarily involved unblinded participants and clinicians,^4,5,24^ creating the possibility that post-randomisation management, surveillance intensity, additional procedures, or crossover between treatment strategies may have differed between treatment groups. However, both trials employed intention-to-treat analyses and demonstrated robust follow-up procedures, limiting the likelihood that such deviations materially influenced mortality estimates.

Three studies were assessed as being at high risk of bias overall. Darke et al. was judged to have some concerns regarding the randomisation process and a high risk of bias in outcome measurement, particularly for graft patency outcomes;^30^ however, the outcomes relevant to the present review, namely mortality and major amputation, were considered less susceptible to measurement bias. Kapfer et al. was judged to have some concerns regarding deviations from allocated treatment and a high risk of bias due to missing outcome data, the latter being directly relevant to the outcomes synthesised in this review.^31^ Sarac et al. was assessed as having a high risk of bias in the randomisation process and some concerns regarding deviations from intended interventions.^29^ Although these limitations may have affected the internal validity and generalisability of the original trial findings, their impact on the pooled estimates generated in the present review is likely to be reduced because outcomes were synthesised within treatment groups rather than through direct between-group comparisons.

#### Non-Randomised Studies of Infrainguinal Bypass (Table 2A)

**Table 2A.** Newcastle-Ottawa Scale (NOS) assessing the risk of bias of non-randomised studies containing infrainguinal cohorts.

|  | Selection |  |  | Comparability |  |  | Outcome |  |  | Total NOS Score (X/9) | Risk of Bias |
| --- | --- | --- | --- | --- | --- | --- | --- | --- | --- | --- | --- |
|  | Representative of exposed cohort | Selection of the non-exposed cohort | Ascertainment of exposure | Outcome not present at start of study | Controls for most important factor | Controls for additional factor(s) | Assessment of outcome | Was follow-up long enough to detect outcomes? | Adequacy of follow-up of cohorts |  |  |
| Attia 2014 | * | * | / | * | * | * | * | * | * | 8 | Low |
| Dick 2007 | * | * | * | * | * | * | * | * | * | 9 | Low |
| Gloviczki 1994 | / | * | * | * | / | / | * | * | / | 5 | Moderate |
| Mezzetto 2024 | * | * | * | * | * | * | * | * | * | 9 | Low |
| Panayiotopoulos 1997 | / | * | * | * | / | / | * | * | / | 5 | Moderate |
| Robbs 1984 | * | * | * | * | / | / | * | * | / | 6 | Moderate |
| Soderstrom 2008 | * | * | * | * | * | * | * | * | * | 9 | Low |
| Stavroulakis 2018 | * | * | * | * | * | * | * | * | / | 8 | Low |
| Takeji 2018 | * | * | * | * | * | * | * | * | * | 9 | Low |
| Troisi 2022 | * | * | * | * | * | * | * | * | * | 9 | Low |
| Watson 1999 | * | * | * | * | * | / | * | * | * | 8 | Low |
| Wolfe 2000 | * | * | * | * | / | * | * | * | * | 8 | Low |

Out of the twelve non-randomised studies for infrainguinal bypass, nine were deemed low risk of bias, of which the five studies by Dick et al., Mezzeto et al., Soderstrom et al., Takeji et al., and Troisi et al. all scored full points on their risk of bias assessment.^19,22,41,42,44^ Stavroulakis et al. only dropped one star due to the proportion lost to follow-up,^21^ whereas Attia et al. only dropped one star for not providing clear information on how exposure ascertainment was achieved.^45^ Both Watson et al. and Wolfle et al. dropped a star due to issues with comparability.^40,43^

The remaining three studies by Glovickzi et al., Robbs et al., and Panayiotopoulos et al. were all deemed moderate risk of bias. This is because all studies scored zero stars in the comparability domain, as well as losing a further star due to concerns around follow-up reporting.^37–39^ In addition to this, both Glovickzi et al. and Panayiotopoulos et al. lost a star in selection due to the cohort only representing a select subgroup of patients with CLTI amenable to bypass.^38,39^

#### Non-Randomised Studies of Major Lower Limb Amputation (Table 2B)

**Table 2B.** Newcastle-Ottawa Scale (NOS) assessing the risk of bias of non-randomised studies containing major lower limb amputation cohorts.

|  | Selection |  |  | Comparability |  |  | Outcome |  |  | Total NOS Score (X/9) | Risk of Bias |
| --- | --- | --- | --- | --- | --- | --- | --- | --- | --- | --- | --- |
|  | Representative of exposed cohort | Selection of the non-exposed cohort | Ascertainment of exposure | Outcome not present at start of study | Controls for most important factor | Controls for additional factor(s) | Assessment of outcome | Was follow-up long enough to detect outcomes? | Adequacy of follow-up of cohorts |  |  |
| Cosgrove 2002 | * | / | * | * | / | / | * | * | / | 5 | Moderate |
| Eneroth 1992 | * | * | * | * | * | * | * | * | * | 9 | Low |
| Matielo 2008 | * | * | * | * | / | / | * | * | / | 6 | Moderate |
| Ploeg 2005 | * | * | * | * | * | * | * | * | * | 9 | Low |
| Robbs 1984 | * | * | * | * | / | / | * | * | / | 6 | Moderate |
| Torbjoernsson 2022 | / | * | * | * | / | / | * | * | / | 5 | Moderate |
| Wolthius 2006 | / | * | * | * | / | / | * | * | * | 6 | Moderate |

Two out of seven studies were deemed low risk of bias, with both studies by Eneroth et al. and Ploeg et al. scoring maximum stars in all domains.^46,49^ The remaining five studies were deemed moderate risk, all of which scored no stars in the comparability domain.^37,47,48,50,51^ In addition to this, both Robbs et al. and Matielo et al. dropped one star in outcome assessment due to lack of reporting of completeness of follow-up^37,50^. Cosgrove et al., Torbjornsson et al. and Wolthius et al. all dropped one star in the selection domain,^47,48,51^ with Torbjornsson et al. and Cosgrove et al. both also losing a star due to concerns regarding follow-up reporting.^47,51^

### Assessment of Publication Bias (Figure 3)

Funnel plots were generated for outcomes reported by more than 10 studies: 30-day mortality (16 studies), 1-year mortality (12 studies), and 30-day subsequent major amputation (12 studies) following infrainguinal bypass. Visual inspection demonstrated broadly symmetrical distributions around the pooled estimates for all three outcomes, with no clear evidence of publication bias or small-study effects. Greater dispersion was observed among smaller studies, particularly for subsequent amputation outcomes, consistent with the underlying between-study heterogeneity.

**Figure 3.**
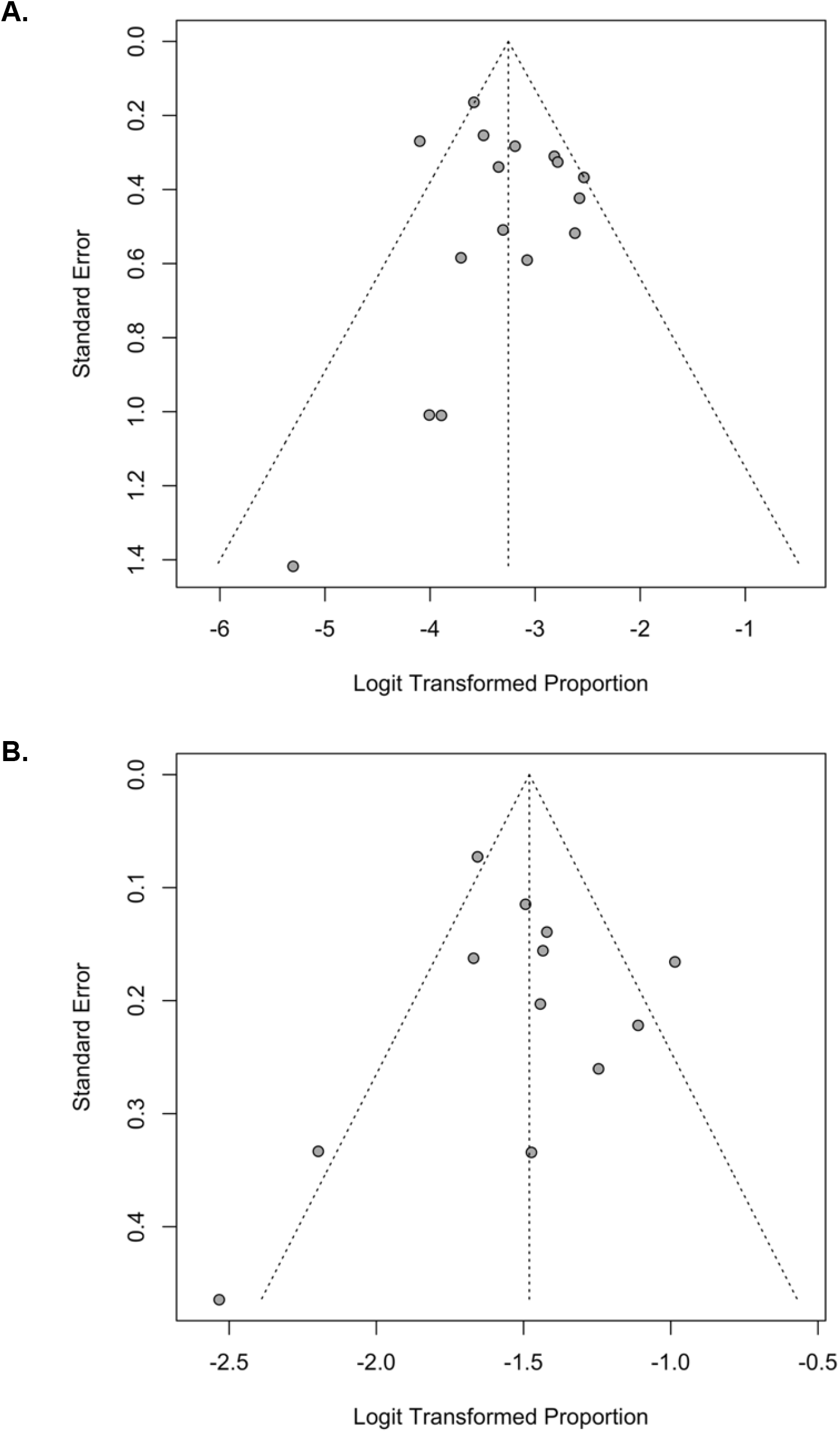
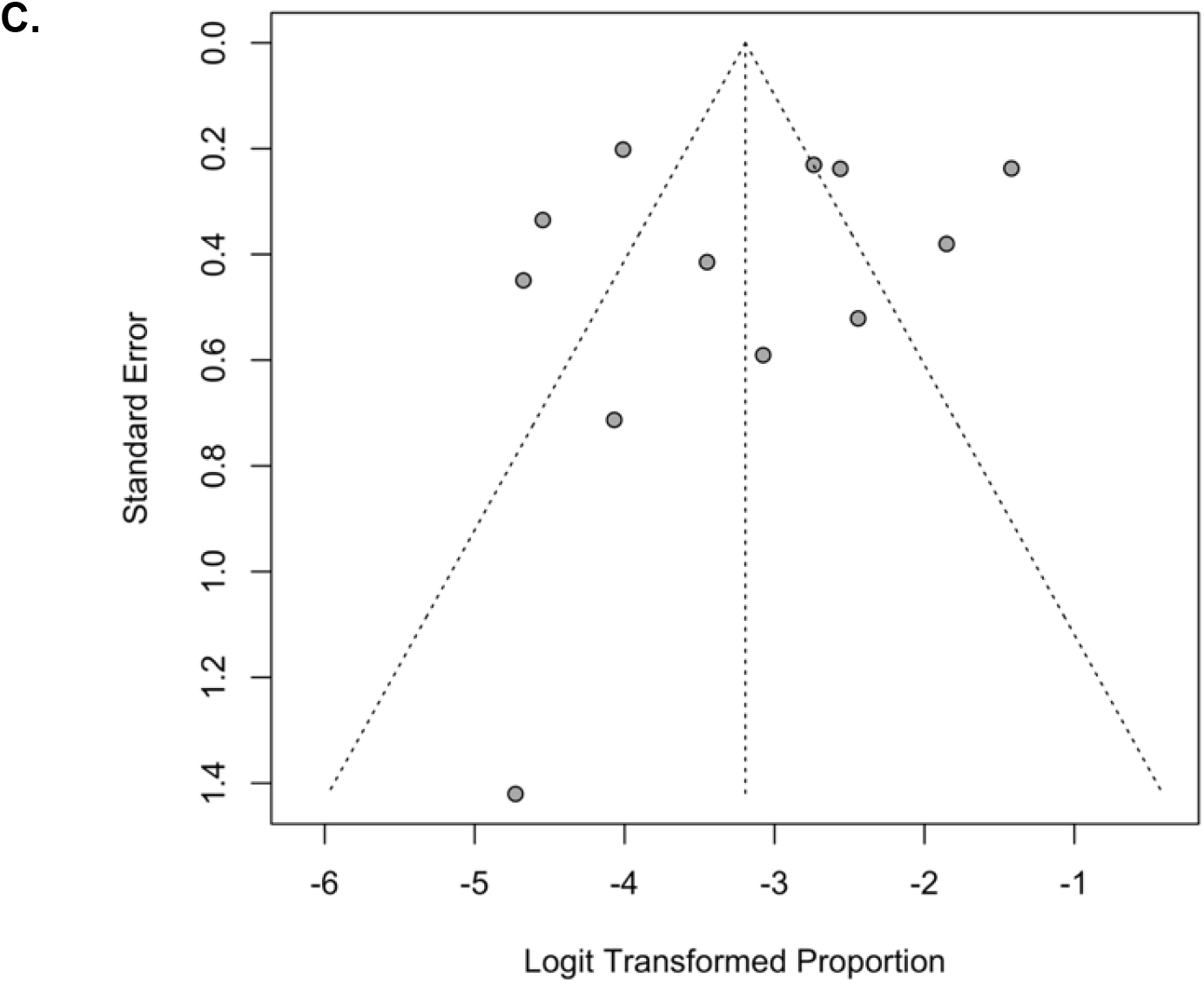
Funnel plot for infrainguinal bypass studies reporting: A) 30-day mortality, B) 1-year mortality, and C) 30-day amputation rate.

### Mortality Outcome (Table 3, Figure 4)

**Figure 4.1.**
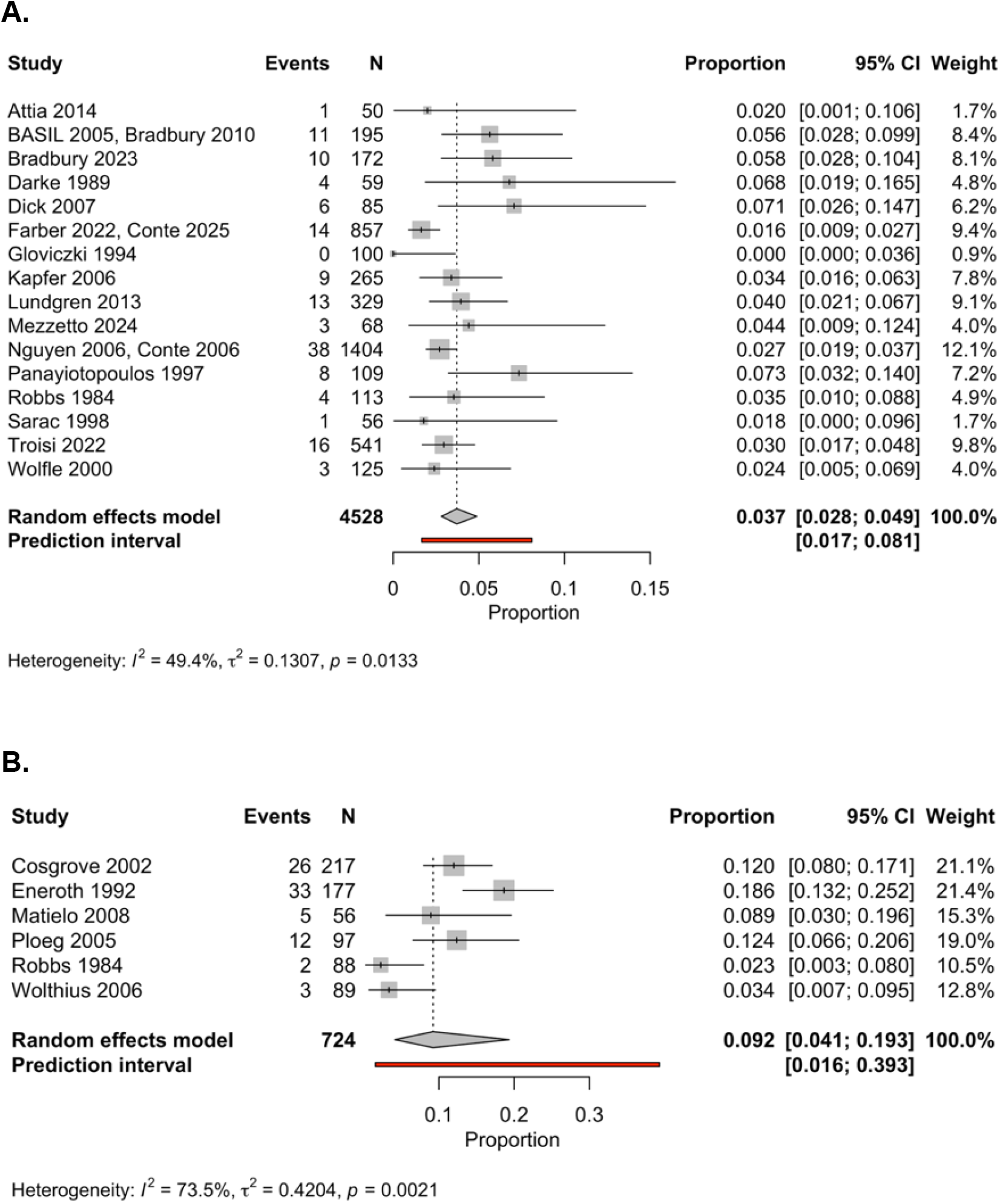
Forest plot summarising the pooled prevalence of 30-day mortality in: A) infrainguinal bypass cohorts, and B) major lower limb amputation cohorts.

**Figure 4.2.**
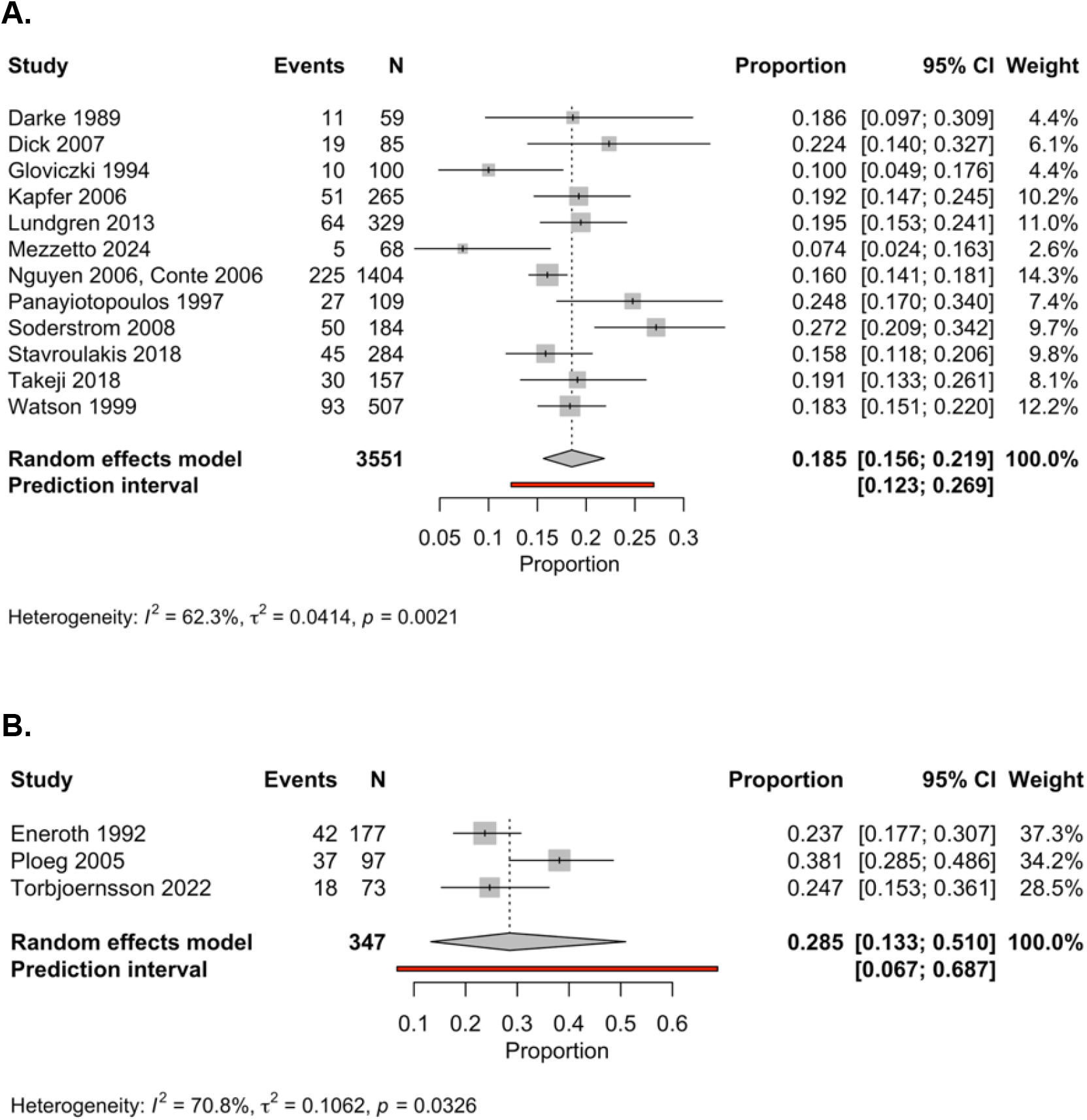
Forest plot summarising the pooled prevalence of 1-year mortality in: A) infrainguinal bypass cohorts, and B) major lower limb amputation cohorts.

**Figure 4.3.**
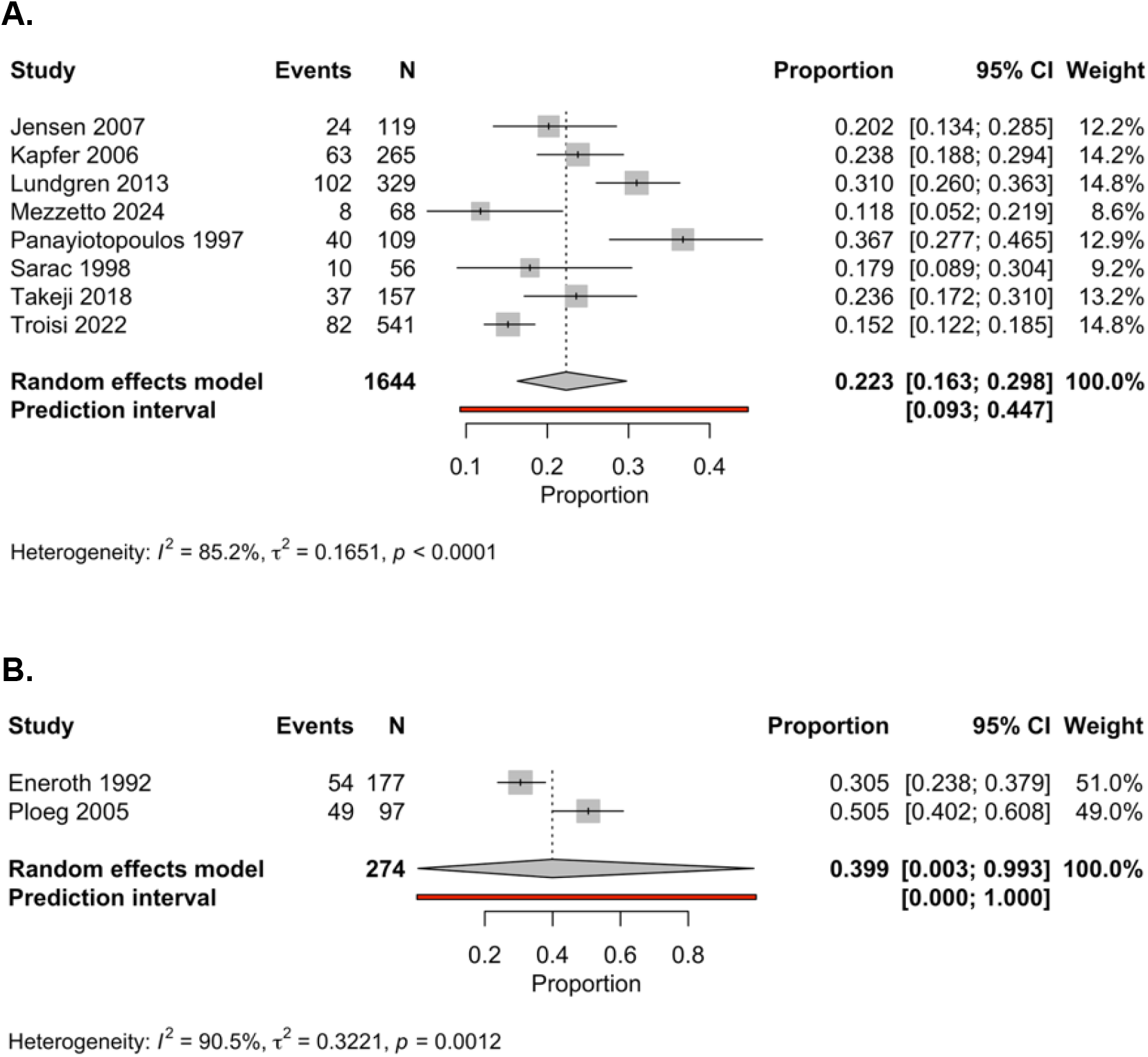
Forest plot summarising the pooled prevalence of 2-year mortality in: A) infrainguinal bypass cohorts, and B) major lower limb amputation cohorts.

**Figure 4.4.**
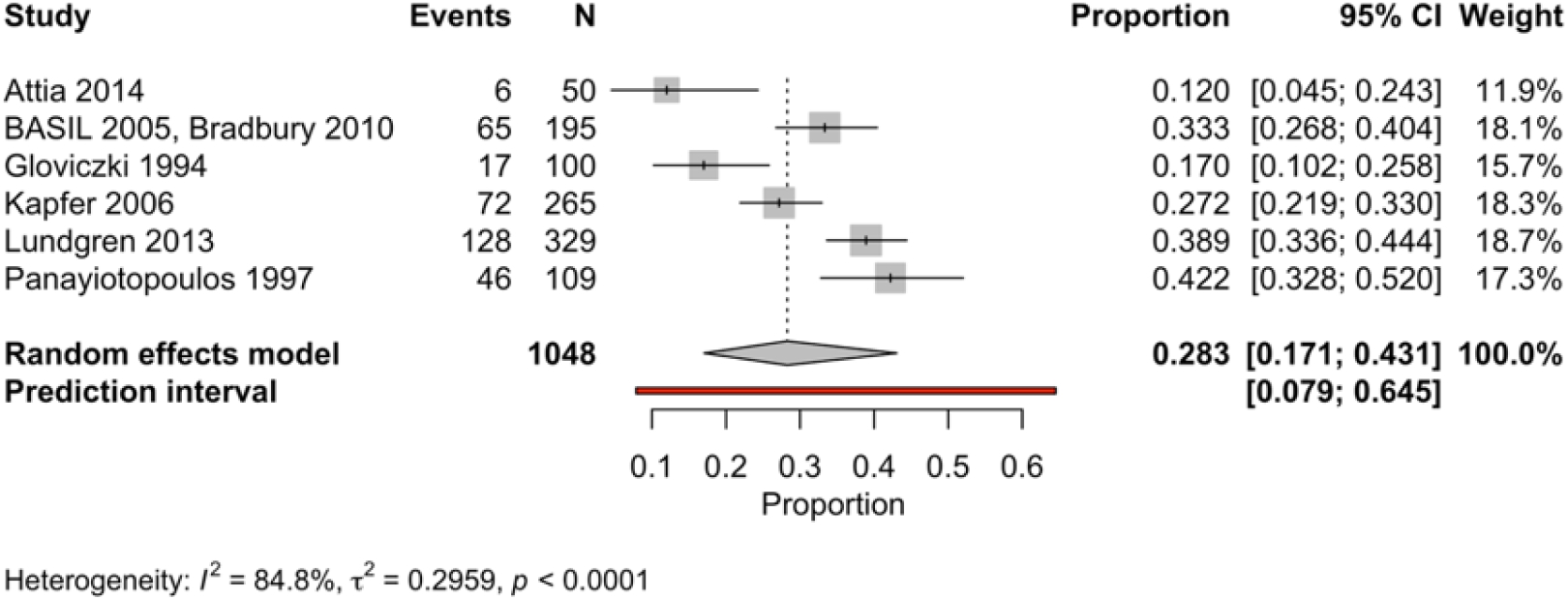
Forest plot summarising the pooled prevalence of 3-year mortality in infrainguinal bypass cohorts. There were no cohorts reporting this outcome for major lower limb amputation.

**Figure 4.5.**
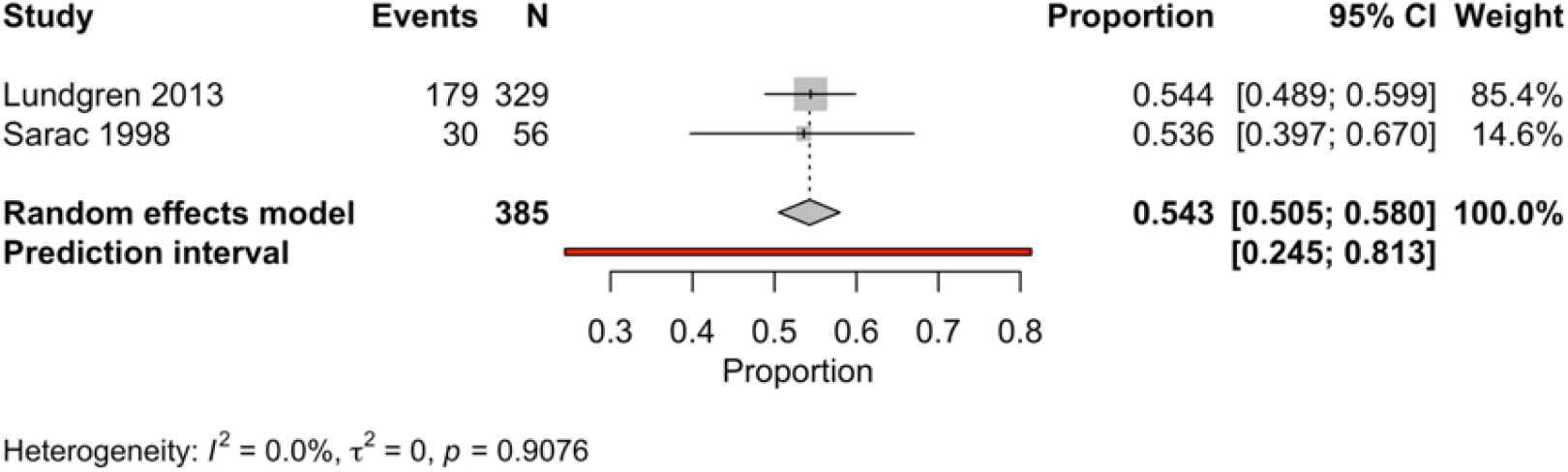
Forest plot summarising the pooled prevalence of 5-year mortality in infrainguinal bypass cohorts. There were no cohorts reporting this outcome for major lower limb amputation.

#### Infrainguinal Bypass

Sixteen studies comprising 4,528 patients reported 30-day mortality following infrainguinal bypass. Individual study mortality estimates ranged from 0% to 7.3%. The pooled 30-day mortality estimate was 3.7% (95% CI 2.8%–4.9%), with moderate between-study heterogeneity (I² = 49.4%), likely reflecting differences in patient selection, anatomical disease burden, operative risk, and underlying healthcare setting.

**Table 3.** Summary of all mortality estimates.

| Timepoint | Infrainguinal Bypass |  |  |  | Major Lower Limb Amputation |  |  |  |
| --- | --- | --- | --- | --- | --- | --- | --- | --- |
|  | Studies | Participants | Estimate (95% CI) | I<sup>2</sup> | Studies | Participants | Estimate (95% CI) | I<sup>2</sup> |
| 30-days | 16 | 4528 | 3.7% (2.8%–4.9%) | 49.4 % | 6 | 724 | 9.2% (4.1%–19.3%) | 73.5 % |
| 1-year | 12 | 3551 | 18.5% (15.6%–21.9%) | 62.3 % | 3 | 347 | 28.5% (13.3%–51.0%) | 70.8 % |
| 2-years | 8 | 1644 | 22.3% (16.3%–29.8%) | 85.2 % | 2 | 274 | 39.9% (0.3%–99.3%) | 90.5 % |
| 3-years | 6 | 1048 | 28.3% (17.1%–43.1%) | 84.8 % | - | - | - | - |
| 5-years | 2 | 385 | 54.3% (50.5%–58.0%) | 0.0% | - | - | - | - |

Mortality increased progressively with longer follow-up. Twelve studies including 3,551 patients contributed 1-year mortality data, with individual estimates ranging from 7.4%^44^ to 27.2%^22^. Overall, pooled mortality at 1 year was 18.5% (95% CI 15.6%–21.9%), with moderate heterogeneity (I² = 62.3%), indicating meaningful variability between study populations.

Eight studies involving 1,644 patients reported mortality at 2 years. Individual study estimates ranged from 11.8%^44^ to 36.7%^38^. The pooled 2-year mortality estimate was 22.3% (95% CI 16.3%–29.8%), with substantial heterogeneity (I² = 85.2%). This heterogeneity likely reflects differences in baseline cardiovascular risk, severity of limb-threatening ischaemia, and competing mortality risks rather than differences in procedural success alone.

At 3 years, six studies including 1,048 patients reported mortality outcomes. Mortality estimates ranged from 12.0%^45^ to 42.2%^38^, with the landmark BASIL trial cohort reporting mortality of 33.3% at 3 years^3,34^. The pooled 3-year mortality estimate was 28.3% (95% CI 17.1%–43.1%), with substantial heterogeneity (I² = 84.8%). These findings demonstrate sustained attrition following bypass, with mortality continuing to rise beyond the immediate perioperative period.

Longer-term mortality was reported by only two studies including 385 patients. Lundgren et al. and Sarac et al. demonstrated remarkably similar 5-year mortality estimates of 54.4% and 53.6%, respectively.^29,33^ The pooled 5-year mortality therefore reached 54.3% (95% CI 50.5%–58.0%), although interpretation is limited by the small number of contributing studies and participants.

#### Major Lower Limb Amputation

Six studies including 724 patients reported 30-day mortality following MLLA. Mortality estimates varied considerably between studies, ranging from 2.3%^37^ to 18.6%.^46^ The remaining studies demonstrated intermediate early mortality. The pooled 30-day mortality following MLLA was 9.2% (95% CI 4.1%–19.3%), more than twice that observed following bypass, but notably there was substantial heterogeneity (I² = 73.5%).

At 1 year, three studies comprising 347 patients contributed mortality data. Individual study estimates ranged from 23.7%^46^ to 38.1%^49^. The pooled 1-year mortality estimate was 28.5% (95% CI 13.3%–51.0%), with substantial heterogeneity (I² = 70.8%).

Two studies including 274 patients reported mortality at 2 years. Ploeg et al. reported mortality of 50.5%,^49^ while Eneroth et al. reported a lower estimate of 30.5%.^46^ The pooled estimate was 39.9% (95% CI 0.3%–99.3%), with very high heterogeneity (I² = 90.5%). The extremely wide confidence interval reflects substantial uncertainty arising from the limited number of available studies and differences between cohorts.

Only one study reported mortality beyond 2-years. Ploeg et al. reported mortality of 71% at 5-years within their cohort.^49^

### Major Adverse Cardiovascular Events (MACE) Outcome (Figure 5)

Six studies including 3,121 patients reported 30-day major adverse cardiovascular events following infrainguinal bypass. Individual study estimates ranged from 4.6% in the BEST-CLI trial^5,24^ to 13.2% by Mezzetto et al.^44^ The pooled 30-day MACE rate of 6.5% (95% CI 4.3%–9.7%), with moderate heterogeneity between studies (I² = 63.5%). The variation observed across cohorts may reflect differences in cardiovascular risk profiles, definitions of cardiovascular outcomes, perioperative surveillance, and the era in which patients were treated.

**Figure 5.**
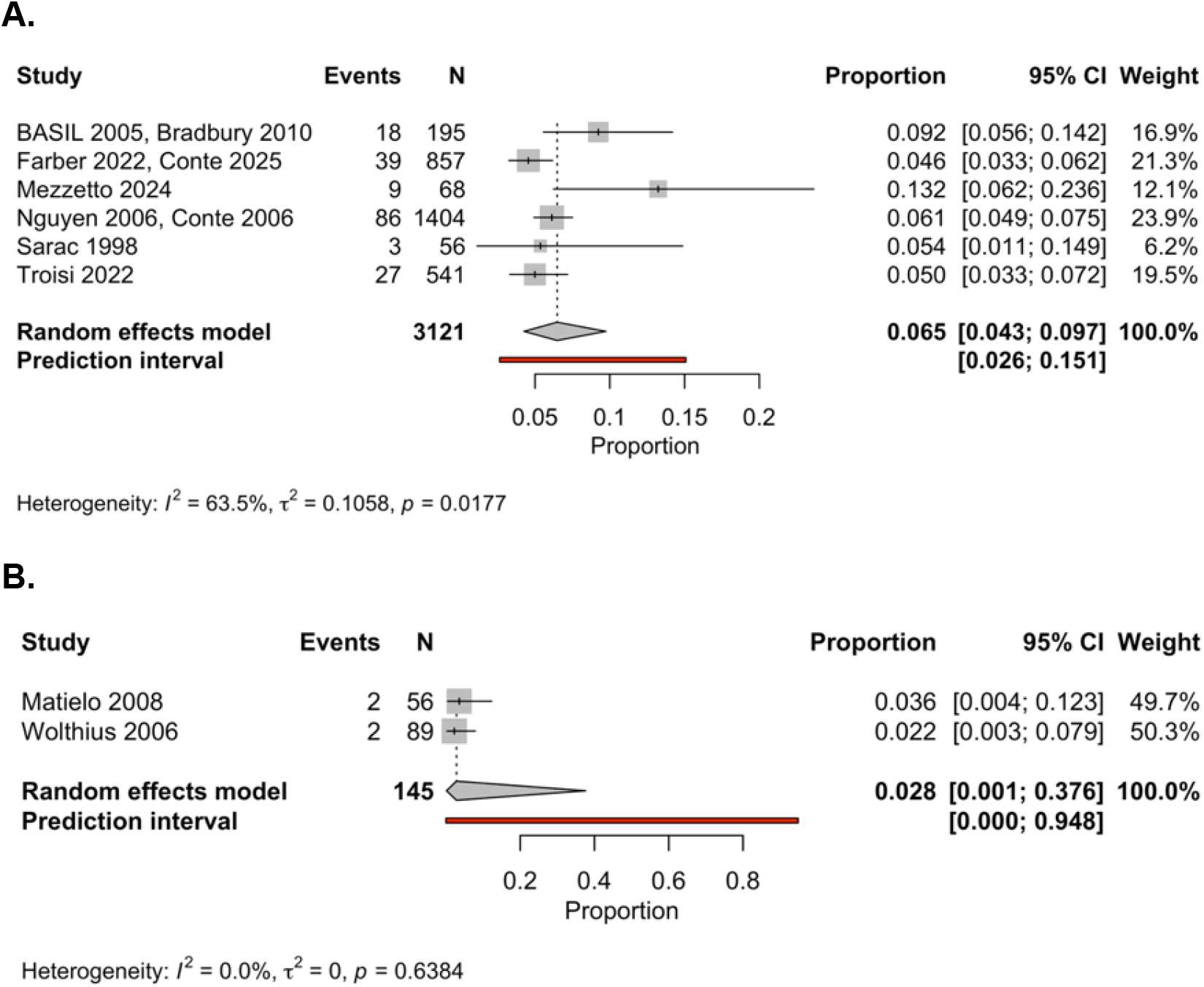
Forest plot summarising the pooled prevalence of 30-day MACE in: A) infrainguinal bypass cohorts, and B) major lower limb amputation cohorts.

Only two studies comprising 145 patients reported 30-day MACE following MLLA, with similar event rates in both these small cohorts. Wolthuis et al. reported a MACE rate of 2.3%^48^, while Matielo et al. reported 3.6%.^50^ The pooled estimate was 2.8% (95% CI 0.1%–37.6%), with no observed statistical heterogeneity (I² = 0%). However, the very wide confidence interval demonstrates substantial imprecision due to the limited number of contributing studies and small overall sample size.

No studies reported MACE outcomes beyond the 30-day postoperative period following either infrainguinal bypass or major lower limb amputation. Therefore, the longer-term cardiovascular burden following these treatment strategies could not be assessed within this review.

### Subsequent Amputation & Re-Amputation Outcome (Table 4, Figure 6)

**Figure 6.1.**
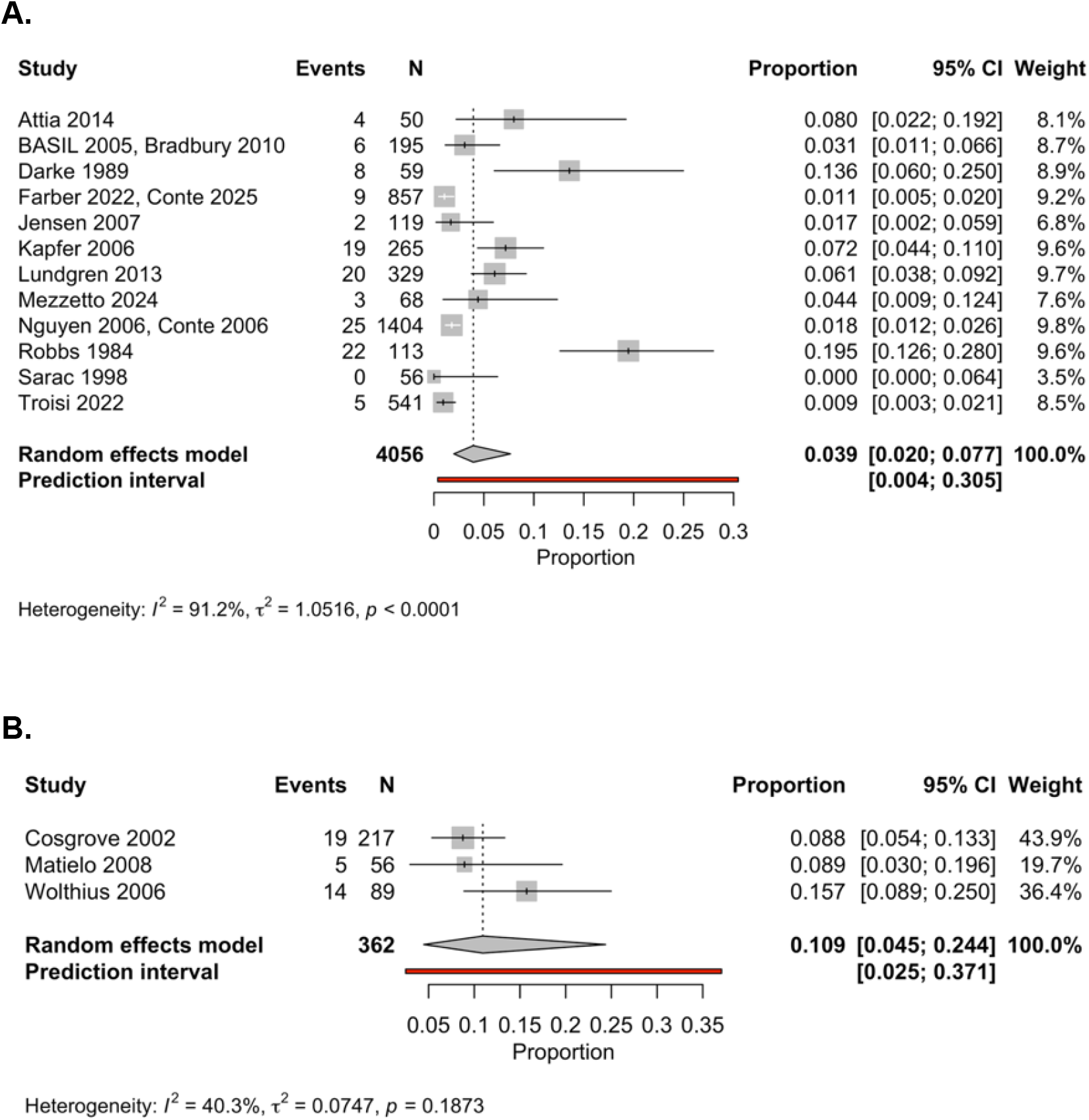
Forest plot summarising the pooled prevalence of 30-day subsequent amputations in: A) infrainguinal bypass cohorts, and B) major lower limb amputation cohorts.

**Figure 6.2.**
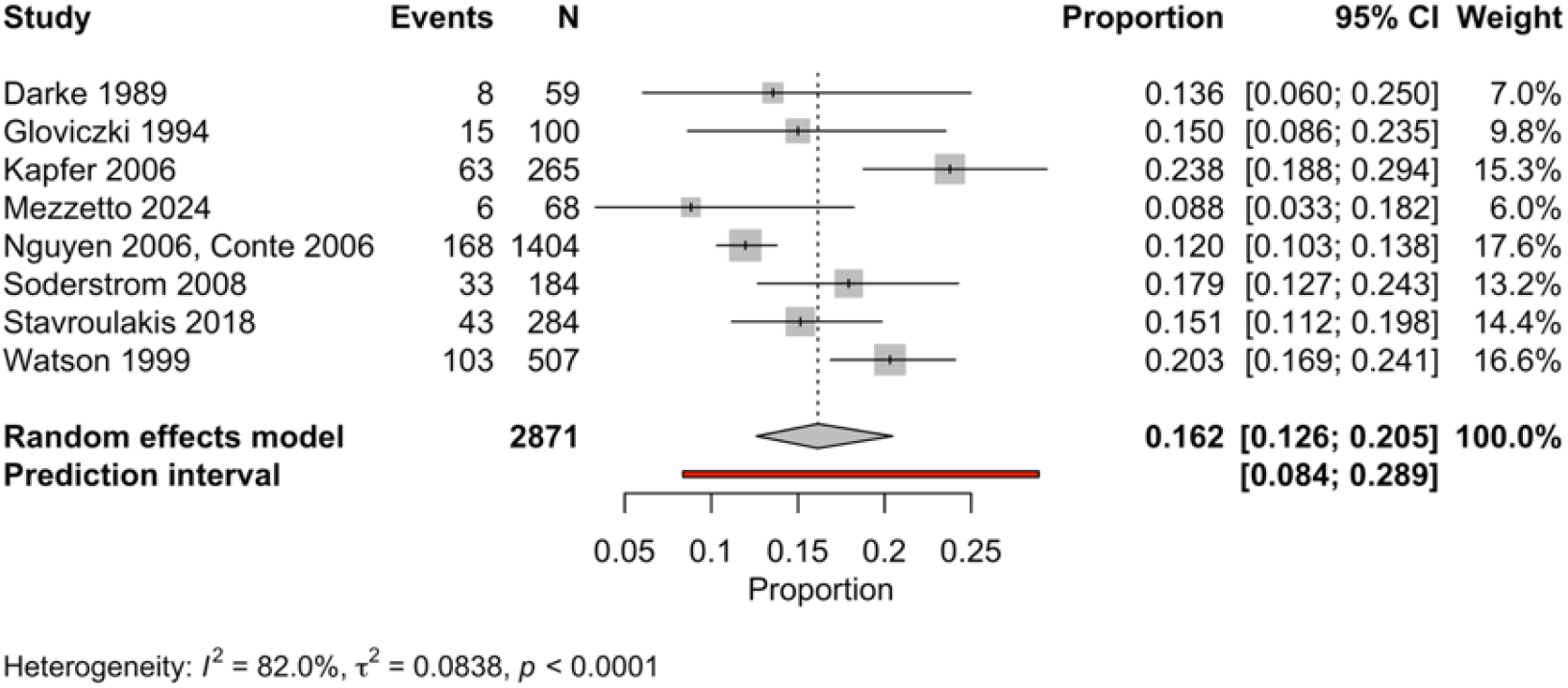
Forest plot summarising the pooled prevalence of 1-year subsequent amputations in infrainguinal bypass cohorts. There were no cohorts reporting this outcome for major lower limb amputation.

**Figure 6.3.**
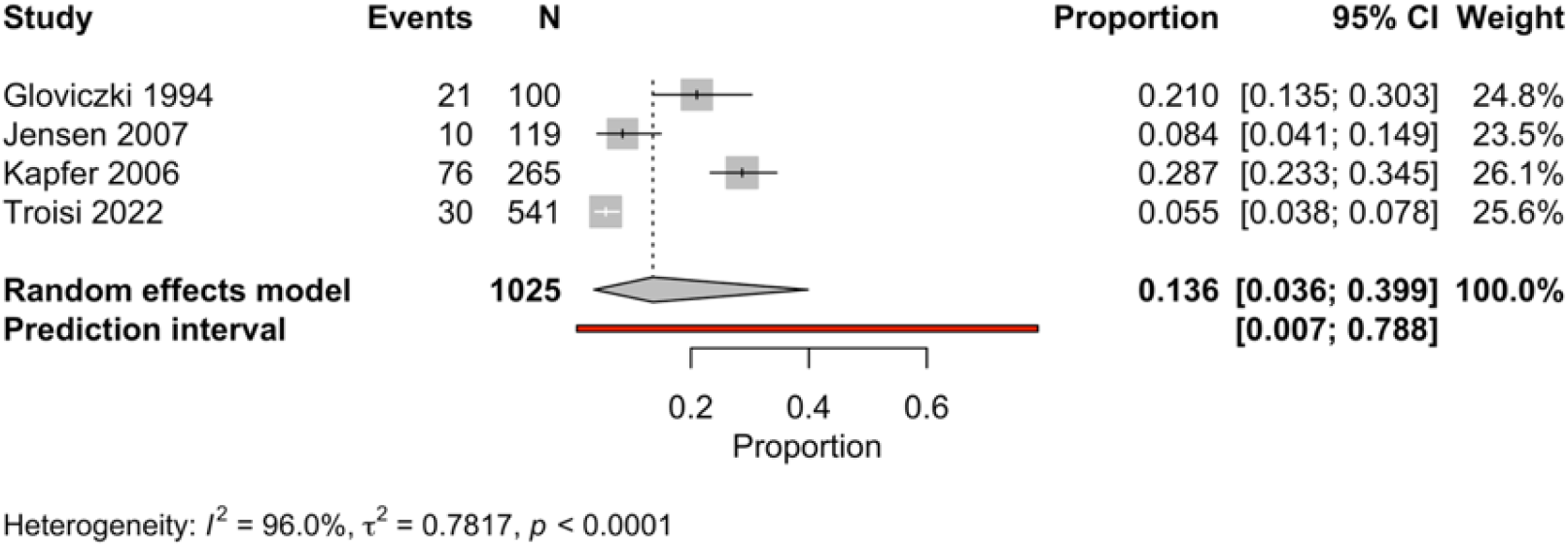
Forest plot summarising the pooled prevalence of 2-year subsequent amputations in infrainguinal bypass cohorts. There were no cohorts reporting this outcome for major lower limb amputation.

**Figure 6.4.**
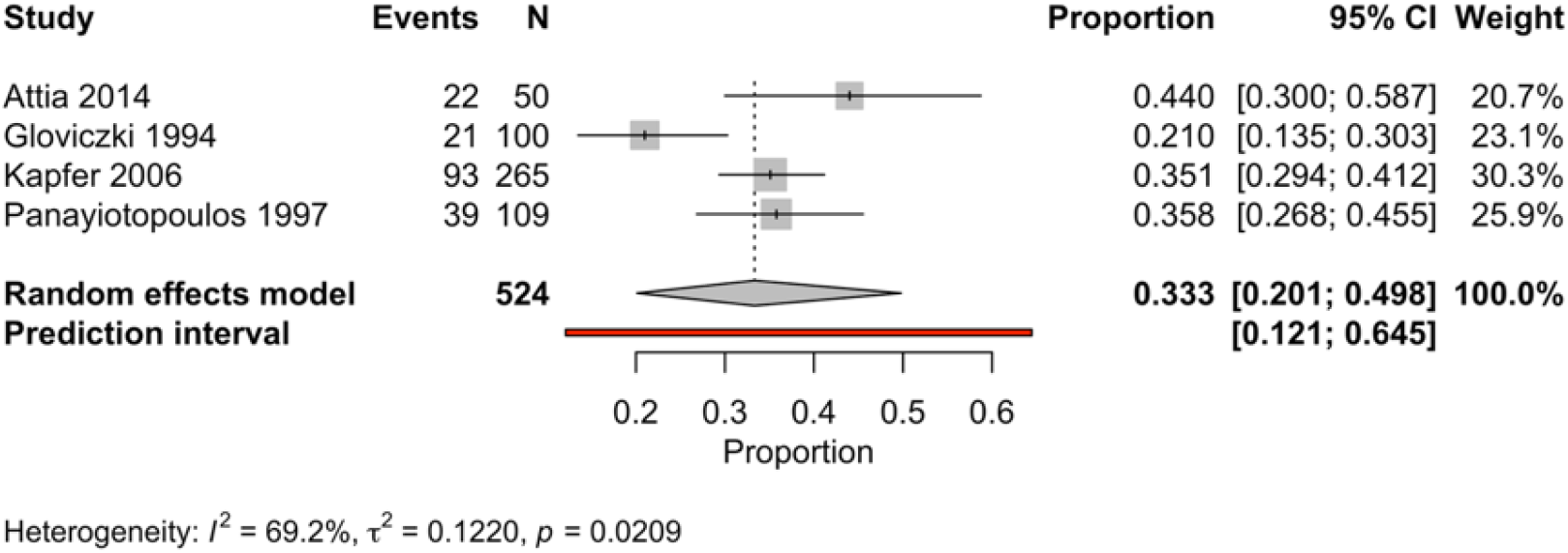
Forest plot summarising the pooled prevalence of 3-year subsequent amputations in infrainguinal bypass cohorts. There were no cohorts reporting this outcome for major lower limb amputation.

#### Infrainguinal Bypass

Twelve studies including 4,056 patients reported subsequent major amputation within 30 days following infrainguinal bypass. Individual study estimates demonstrated substantial variation, ranging from 0% by Sarac et al.^29^ to 19.5% by Robbs et al.^37^ Of note, the larger contemporary cohorts generally reported lower rates, including BEST-CLI (1.1%)^5,24^, LIMBSAVE (0.9%),^19^ and PREVENT-III (1.8%).^35,36^ The pooled 30-day subsequent amputation rate following bypass was therefore 3.9% (95% CI 2.0%–7.7%), with substantial heterogeneity (I² = 91.2%). The marked variability between studies likely reflects differences in disease severity, limb selection, definitions of treatment failure, and thresholds for early amputation following unsuccessful revascularisation.

**Table 4.**
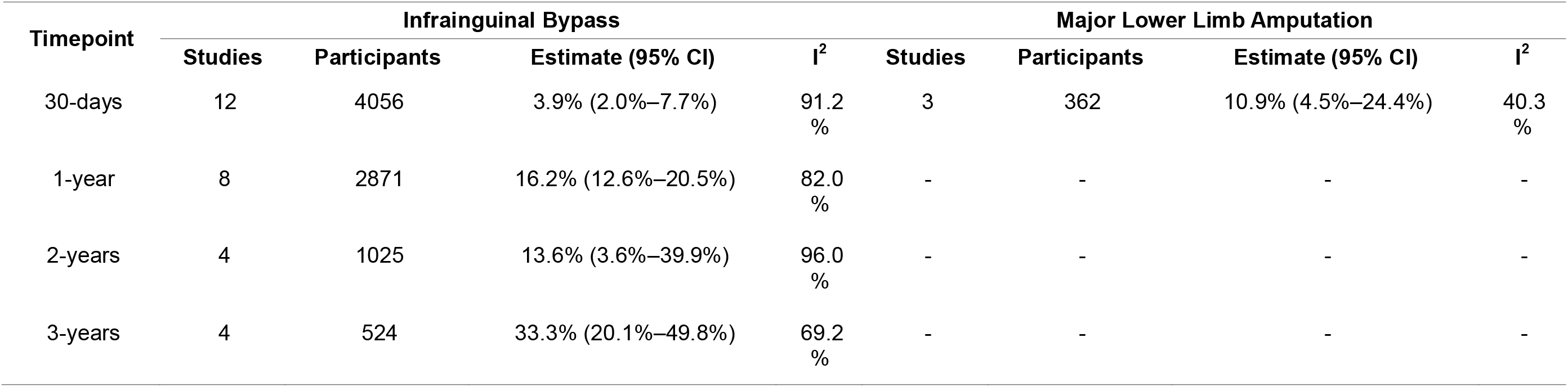
Summary of all (re-)amputation estimates: for infrainguinal bypass, this represents a subsequent amputation following the initial bypass, but for major lower limb amputations, this represents a re-amputation at a higher level than the original procedure.

At 1 year, eight studies including 2,871 patients reported subsequent amputation. Event rates ranged from 8.8%^44^ to 23.8%^31^. Other studies demonstrated relatively consistent intermediate rates, and no correlation with sample size or study period was observed. The pooled 1-year subsequent amputation rate was 16.2% (95% CI 12.6%–20.5%), with substantial heterogeneity (I² = 82.0%).

Four studies including 1,025 patients contributed data at 2 years. Individual estimates varied considerably, ranging from 5.6%^19^ to 28.7%^31^. The pooled estimate was 13.6% (95% CI 3.6%–39.9%), with very high heterogeneity (I² = 96.0%). The wide confidence interval and extreme heterogeneity indicate considerable uncertainty and suggest that long-term limb outcomes following bypass vary substantially between these patient cohorts.

At 3 years, four studies comprising 524 patients reported subsequent amputation. Rates ranged from 21.0% in Gloviczki et al.^39^ to 44.0% in Attia et al.^45^ The pooled 3-year estimate was 33.3% (95% CI 20.1%–49.8%), demonstrating that despite initial limb salvage following revascularisation, a substantial proportion of patients ultimately progress to major amputation over longer follow-up.

#### Major Lower Limb Amputation

Three studies including 362 patients reported subsequent amputation events within 30 days following an initial major lower limb amputation. Individual study estimates ranged from 8.9% in Matielo et al.^50^ to 15.7% in Wolthuis et al.^48^ The pooled estimate was therefore 10.9% (95% CI 4.5%–24.4%), with moderate heterogeneity (I² = 40.3%).

No studies reported longer-term re-amputation outcomes. Therefore, the available evidence only permits assessment of early postoperative revision or further amputation events.

### Baseline Co-Morbidities and Health (Table 5; Supplementary Material 2)

Baseline co-morbidities were common in both treatment groups, reflecting the substantial cardiovascular disease burden associated with CLTI. Considerable heterogeneity was observed across studies for most baseline characteristics.

**Table 5.**
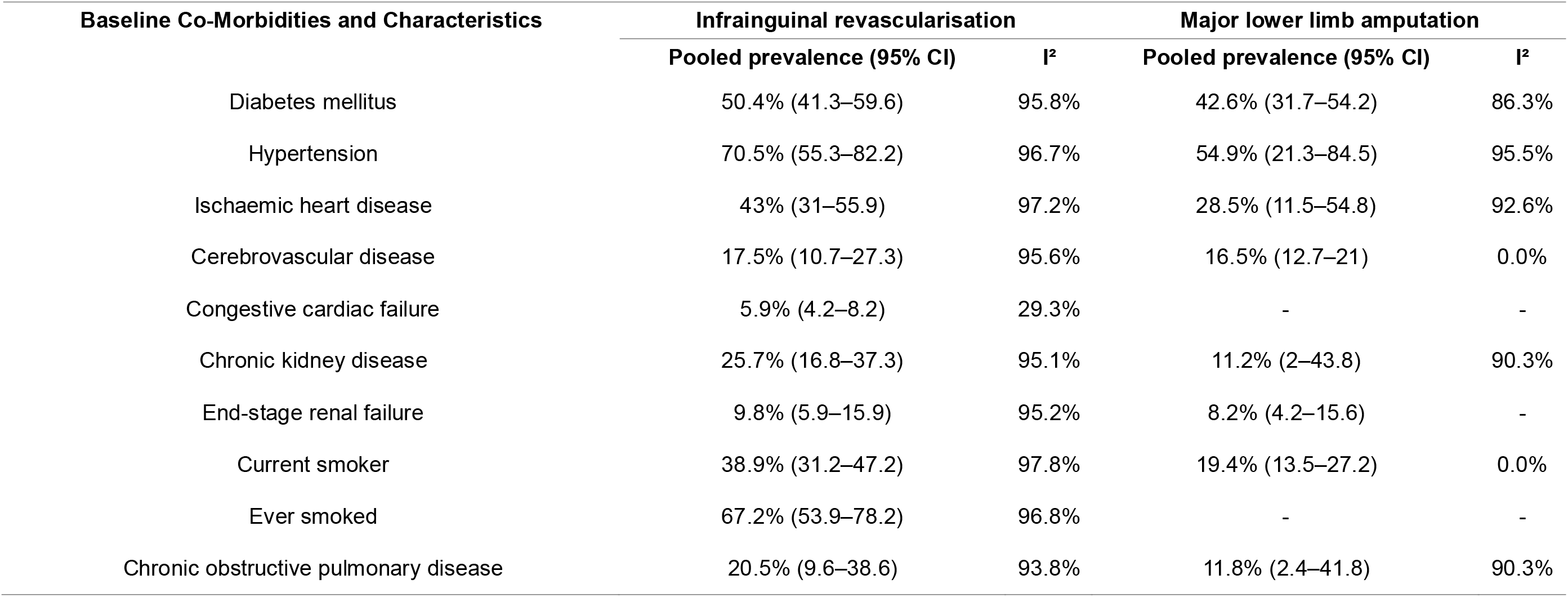
Baseline characteristics summary of all studies included within the review. Pooled prevalence presented using random effect models due to significant heterogeneity across the studies.

#### Infrainguinal Bypass

Among patients undergoing infrainguinal bypass, hypertension was the most prevalent co-morbidity, affecting 70.5% of patients (95% CI, 55.3%–82.2%; I² = 96.7%). Diabetes mellitus was present in approximately half of all patients (50.4%; 95% CI, 41.3%–59.6%; I² = 95.8%). Ischaemic heart disease affected 43.0% of patients (95% CI, 31.0%–55.9%; I² = 97.2%), while cerebrovascular disease was present in 17.5% (95% CI, 10.7%–27.3%; I² = 95.6%).

Renal impairment was also common. Chronic kidney disease was reported in 25.7% of patients (95% CI, 16.8%–37.3%; I² = 95.1%), while end-stage renal failure was present in 9.8% (95% CI, 5.9%–15.9%; I² = 95.2%). Congestive cardiac failure was less frequently reported, with a pooled prevalence of 5.9% (95% CI, 4.2%–8.2%; I² = 29.3%).

Smoking exposure was highly prevalent. Current smoking was reported in 38.9% of patients (95% CI, 31.2%–47.2%; I² = 97.8%), while a history of smoking was reported in 67.2% (95% CI, 53.9%–78.2%; I² = 96.8%). Chronic obstructive pulmonary disease was present in 20.5% of patients (95% CI, 9.6%–38.6%; I² = 93.8%).

#### Major Lower Limb Amputation

Among patients undergoing major lower limb amputation, hypertension remained the most common co-morbidity, with a pooled prevalence of 54.9% (95% CI, 21.3%–84.5%; I² = 95.5%). Diabetes mellitus affected 42.6% of patients (95% CI, 31.7%–54.2%; I² = 86.3%), while ischaemic heart disease was reported in 28.5% (95% CI, 11.5%–54.8%; I² = 92.6%).

Cerebrovascular disease was present in 16.5% of patients (95% CI, 12.7%–21.0%; I² = 0.0%), representing one of the few characteristics with minimal between-study heterogeneity. Chronic kidney disease was reported in 11.2% of patients (95% CI, 2.0%–43.8%; I² = 90.3%), and end-stage renal failure in 8.2% (95% CI, 4.2%–15.6%). Chronic obstructive pulmonary disease affected 11.8% of patients (95% CI, 2.4%–41.8%; I² = 90.3%).

Current smoking was reported in 19.4% of patients (95% CI, 13.5%–27.2%; I² = 0.0%). Data regarding previous smoking history and congestive cardiac failure were insufficient for pooled analysis in the amputation cohorts.

## Discussion

This is the first systematic review and meta-analysis to date that assesses mortality proportions in both infrainguinal bypass and MLLA cohorts for CLTI. The Global Vascular Guidelines (GVG) emphasise that management should be driven by the PLAN framework (patient risk, limb severity, and anatomic complexity) and that primary amputation is appropriate only for selected patients with an unsalvageable limb, poor function, or limited life expectancy after shared decision-making.^1^ Our findings fit that framework closely, but they also expose a major weakness in the current evidence base: the literature is far richer for bypass than for amputation, while the patients facing these decisions are frequently those in whom the greatest uncertainty remains.

We found that several important differences between the bypass and amputation evidence bases were evident. First, substantially more evidence was available for infrainguinal bypass, both in terms of study number and sample size, with bypass studies accounting for more than four-fifths of all included patients. Second, all randomised evidence identified within the review related to revascularisation, whereas no randomised trials reporting mortality proportions following MLLA were identified. Third, contemporary multicentre and international collaborations were common within the bypass literature but largely absent from the amputation literature. Finally, disease severity reporting was more comprehensive among bypass studies, whereas amputation cohorts were generally characterised by broader descriptions of gangrene, tissue loss and advanced CLTI. These findings highlight the substantial imbalance in the quantity and quality of evidence available to inform decision-making between limb salvage and MLLA.

Infrainguinal bypass is associated with relatively low early mortality, but survival reduces significantly as time progresses. Approximately one in five patients died within the first year following bypass, increasing to more than half by 5 years. In comparison, across all reported timepoints, mortality following MLLA remained consistently high, with approximately one in ten patients dead within 30 days, increasing to almost one-third by 1 year and approximately two-fifths by 2 years. Compared with bypass cohorts, mortality appeared consistently higher after amputation, although direct comparison is limited by substantial differences in baseline patient characteristics and clinical indications for treatment. The practical message here remains that neither strategy is risk-free, and neither should be framed as a simple fix for a significant health problem.

The limb outcomes are just as important as the mortality outcomes, because in CLTI the goal is not merely to keep the patient alive but to preserve a functional limb that can support ambulation, independence, and wound healing.^1^ After bypass, early major amputation was uncommon, but later failure accumulated with one in three requiring MLLA at 3-years post-bypass, showing that technical success in the operating theatre does not necessarily translate into durable limb salvage over time. This matters in practice: a bypass that buys a few months but is followed by repeated wound failure, revision procedures, or ultimate amputation may still carry a very large burden for the patient. Primary amputation was also not a definitive end point for all patients, with meaningful early re-amputation or revision in some cohorts. Inevitably, the clinical decision making in the surgical management of CLTI is a trade-off between two pathways, each of which can lead to repeated interventions and ongoing disability.

The baseline comorbidity pattern is also highly informative. In this review, bypass cohorts were not simply healthier patients with fewer systemic risk factors; they still carried a heavy burden of diabetes, hypertension, ischaemic heart disease, renal disease, and smoking exposure, with either equivocal or greater pooled prevalences than the MLLA cohorts across all baseline health characteristics. This is important because it argues against a simplistic interpretation that better bypass outcomes are explained only by “fitter” patients. However, one of the major limitations here is that the amputation literature, by contrast, is smaller and less granular, with less consistent reporting of disease severity and comorbidity, which makes apparent differences in baseline profiles difficult to interpret as true biological differences rather than differences in selection, recording, and referral pathways. Future research into the health profiles of patients that receive these different treatments, and thus inevitably those profiles that would benefit from one approach over another, is critical to ensure that tailored decisions in CLTI management can be advanced.

The magnitude of heterogeneity across all outcomes studied within this review is also significant. It tells us that CLTI is a syndrome with very different phenotypes, and that studies often mix patients with severe tissue loss, infection, renal dysfunction, frailty, and different distal targets or conduit options. The GVG explicitly notes that the field has been hampered by a lack of uniform staging and an incomplete picture of patient-focused outcomes, ^1^ and that remains true here. Mortality was the most consistently captured endpoint, but it is not the only one that matters. Function, wound healing, quality of life, mobility, and treatment burden are inconsistently reported, leaving an evidence base that is rich in survival numbers but thin in what patients actually experience.

These findings have immediate implications for practice. First, bypass should not be offered or defended on the basis of limb salvage alone. The more honest framing is that bypass is a limb-preserving strategy that may be appropriate when there is a realistic chance of functional recovery and meaningful life expectancy, but it carries a substantial long-term mortality burden because the underlying disease remains active. Second, primary amputation should not be presented as the “safer” option by default in those with unsalvageable limbs, poor functional status, or short life expectancy, because mortality remains high and revision is not rare. Third, the real clinical decision is often not bypass versus amputation, but rather which treatment offers the best chance of the outcome the patient values most: pain control, ambulation, time at home, avoidance of repeated procedures, or simply the least burdensome path through a terminal vascular syndrome.

The paper also highlights where the field still falls short. There are no randomised data for primary amputation, very limited longer-term cardiovascular outcome reporting, and almost no consistent reporting of patient-reported outcomes. Risk stratification was also underused in the older literature, but this may improve in the future given that WIfI staging is now central to CLTI assessment and is specifically linked to patient-centred outcomes in contemporary vascular practice. Future comparative work should therefore prioritise prospective cohorts and target-trial style analyses that capture not only survival, but also function, quality of life, wound healing, rehospitalisation, and the real burden of recovery. Without those data, decision-making will continue to rely on incomplete proxies for benefit and harm.

## Conclusion

This review shows that CLTI is a high-mortality systemic disease in which both bypass and primary major amputation carry substantial downstream risk. Bypass offers better early limb preservation and lower early mortality, but long-term survival remains poor and later limb failure is common. Primary amputation remains an important option for selected patients, but it is not a low-risk fallback. The most important conclusion is therefore not that one procedure “wins,” but that treatment should be individualised, multidisciplinary, and grounded in honest discussion of prognosis, function, and patient priorities.

## Data Availability

All data produced in the present study are available upon reasonable request to the authors.

## Acknowledgements

Jordan Green is funded through a NIHR Academic Clinical Fellowship. Henry Davies is funded through a NIHR Clinical Lectureship. David Russell is supported in part by the National Institute for Health and Care Research (NIHR) Leeds Biomedical Research Centre (BRC) (NIHR203331) and through an NIHR Advanced Fellowship (NIHR300633). The views expressed are those of the author(s) and not necessarily those of the NHS, the NIHR, or the Department of Health and Social Care.

