## Supplementary Material 1 for "Revascularisation versus amputation for chronic limb-threatening ischaemia: a systematic review and meta-analysis of clinical outcomes and patient characteristics"

Headings, key words and phrases used as part of the literature search for each database.

|  | **Medline** | **Embase** | **Cinahl** | **Cochrane** |
| --- | --- | --- | --- | --- |
| **Critical limb ischaemia** | *Headings:*  exp Peripheral Arterial Disease/  exp Arterial Occlusive Diseases/  Peripheral Vascular Diseases/  exp Chronic Limb-Threatening Ischemia/  *Keywords and phrases:*  critical limb isch*  chronic limb isch*  CLTI  CLI | *Headings:*  exp peripheral occlusive artery disease/  exp peripheral vascular diseases/  limb ischemia/  exp chronic limb threatening ischemia/  exp critical limb ischemia/  *Keywords and phrases:*  critical limb isch*  chronic limb isch*  CLTI  CLI | *Headings:*  (MH “Peripheral Vascular Diseases+”)  (MH “Arterial Occlusive Diseases+”)  (MH “Chronic Limb-Threatening Ischemia”)  *Keywords and phrases:*  critical limb isch*  chronic limb isch*  CLTI  CLI  e.g. (TI “critical limb isch*” or AB “critical limb isch*”) | *Headings:*  exp peripheral arterial disease/  exp arterial occlusive diseases/  exp peripheral vascular diseases/  *Keywords and phrases:*  critical limb isch*  chronic limb isch*  CLTI  CLI |
| **Foot ulceration** | *Headings:*  exp Diabetic Foot/  exp Leg Ulcer/  exp Ulcer/  *Keywords and phrases:*  diabetic foot ulcer  diabetic foot  DFU  foot ulcer  leg wound  foot wound  leg ulcer  isch* ulcer | *Headings:*  exp diabetic foot/  exp leg ulcer/  exp ulcer/  *Keywords and phrases:*  diabetic foot ulcer  diabetic foot  DFU  foot ulcer  leg wound  foot wound  leg ulcer  isch* ulcer | *Headings:*  (MH "Diabetic Foot")  (MH "Leg Ulcer+")  *Keywords and phrases:*  diabetic foot ulcer  diabetic foot  DFU  foot ulcer  leg wound  foot wound  leg ulcer  isch* ulcer | *Headings:*  exp diabetic foot/  exp Leg Ulcer/  exp ulcer/  *Keywords and phrases:*  diabetic foot ulcer  diabetic foot  DFU  foot ulcer  leg wound  foot wound  leg ulcer  isch* ulcer |
| **Infrainguinal bypass** | *Keywords and phrases:*  infrainguinal  bypass  bypass graft  vein graft  prosthetic graft  PTFE graft  Dacron graft | *Headings:*  bypass surgery/  *Keywords and phrases:*  infrainguinal  bypass  bypass graft  vein graft  prosthetic graft  PTFE graft  Dacron graft | *Keywords and phrases:*  infrainguinal  bypass  bypass graft  vein graft  prosthetic graft  PTFE graft  Dacron graft | *Keywords and phrases:*  infrainguinal  bypass  bypass graft  vein graft  prosthetic graft  PTFE graft  Dacron graft |
| **Amputation** | *Headings:*  exp Amputation/  ankle/ or knee/ or leg/ or thigh/ or hip/  *Keywords and phrases:*  major amputation  lower limb amputation  lower extremity amputation  BKA  AKA  TKA  below knee amputation  through knee amputation  above knee amputation  knee disarticulation  Gritti Stokes | *Headings:*  exp leg amputation/  exp leg/  ankle/ or knee/ or thigh/ or hip/  *Keywords and phrases:*  major amputation  lower limb amputation  lower extremity amputation  BKA  AKA  TKA  below knee amputation  through knee amputation  above knee amputation  knee disarticulation  Gritti Stokes | *Headings:*  (MH "Amputation+")  *Keywords and phrases:*  major amputation  lower limb amputation  lower extremity amputation  BKA  AKA  TKA  below knee amputation  through knee amputation  above knee amputation  knee disarticulation  Gritti Stokes | *Headings:*  exp Amputation/  ankle/ or knee/ or leg/ or thigh/ or hip/  *Keywords and phrases:*  major amputation  lower limb amputation  lower extremity amputation  BKA  AKA  TKA  below knee amputation  through knee amputation  above knee amputation  knee disarticulation  Gritti Stokes |
| **Article type** | *Headings:*  exp Evidence-Based Medicine/  exp "Systematic Review"/  exp Meta-Analysis/  exp Controlled Clinical Trials as Topic/  exp Randomized Controlled Trials as Topic/  exp Cohort Studies/  exp Prospective Studies/  exp Clinical Trial/  *Keywords and phrases:*  controlled study  Latin square  clinical trial  randomi* controlled trial  RCT  control* trial  cohort  registry | *Headings:*  exp evidence based medicine/  systematic review/  exp meta analysis/  exp controlled study/  exp randomized controlled trial/  exp cohort analysis/  exp prospective study/  exp clinical trial/  *Keywords and phrases:*  controlled study  Latin square  clinical trial  randomi* controlled trial  RCT  control* trial  cohort  registry | *Headings:*  (MH "Professional Practice, Evidence-Based+")  (MH "Literature Review+")  (MH "Meta Analysis")  (MH "Clinical Trials+")  (MH "Nonexperimental Studies+")  *Keywords and phrases:*  controlled study  Latin square  clinical trial  randomi* controlled trial  RCT  control* trial  cohort  registry |  |
| **Human** | Humans/ | exp human/ | (MH "Human") | Humans/ |
