## Supplementary Material 2 for "Revascularisation versus amputation for chronic limb-threatening ischaemia: a systematic review and meta-analysis of clinical outcomes and patient characteristics"

**SM2.1** Forest plot summarising the pooled prevalence of diabetes mellitus in: A) infrainguinal bypass cohorts, and B) major lower limb amputation cohorts.

**A.**

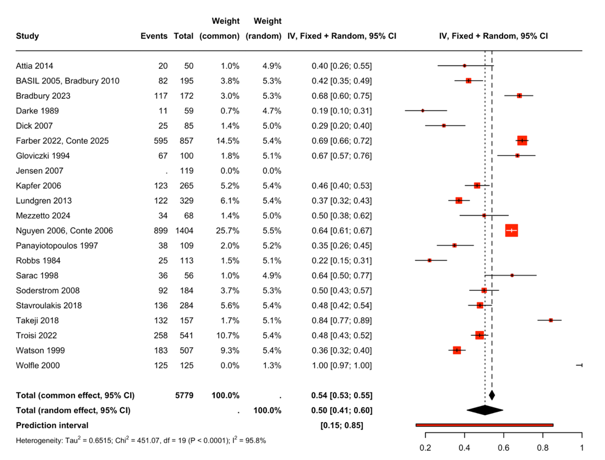

**B.**

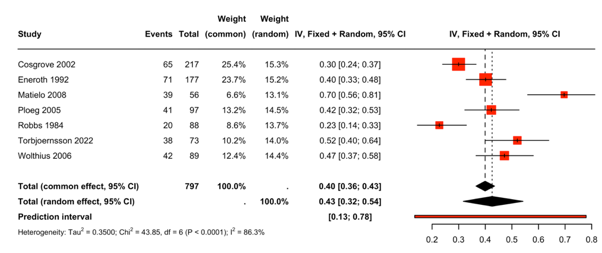

**SM2.2** Forest plot summarising the pooled prevalence of hypertension in: A) infrainguinal bypass cohorts, and B) major lower limb amputation cohorts.

**A.**

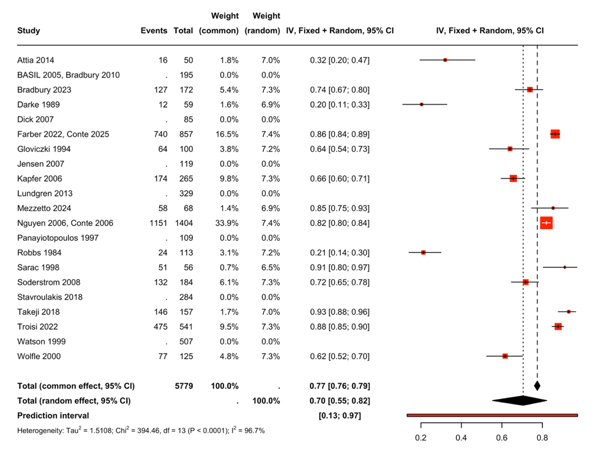

**B.**

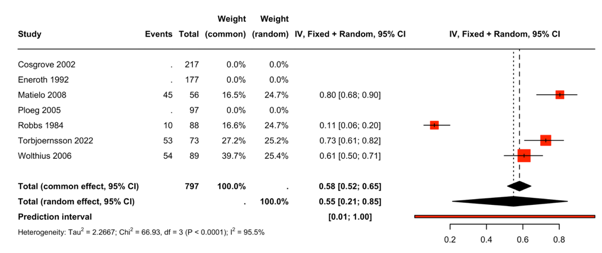

**SM2.3** Forest plot summarising the pooled prevalence of ischaemic heart disease in: A) infrainguinal bypass cohorts, and B) major lower limb amputation cohorts.

**A.**

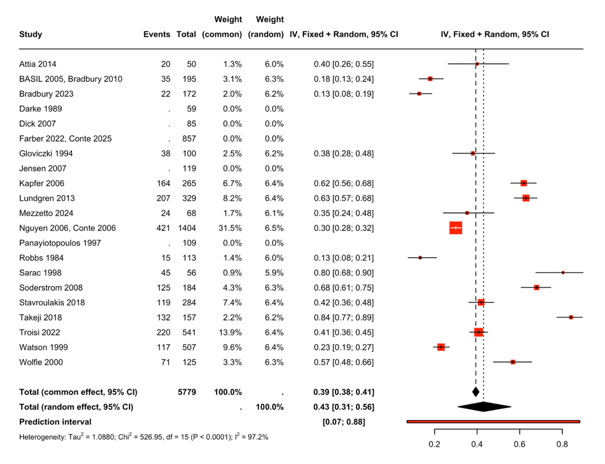

**B.**

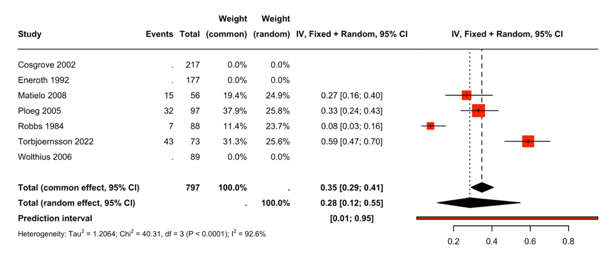

**SM2.4** Forest plot summarising the pooled prevalence of cerebrovascular disease in: A) infrainguinal bypass cohorts, and B) major lower limb amputation cohorts.

**A.**

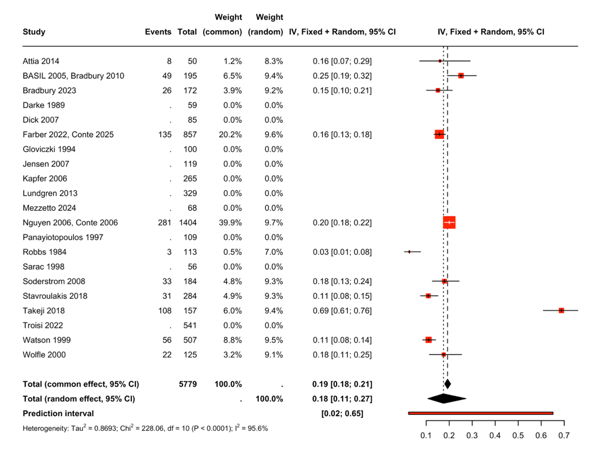

**B.**

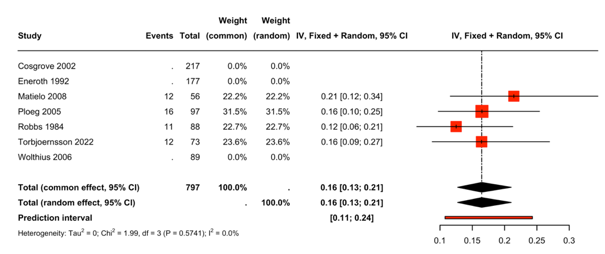

**SM2.5** Forest plot summarising the pooled prevalence of congestive cardiac failure in infrainguinal bypass cohorts. There were no studies reporting this for major lower limb amputation cohorts.

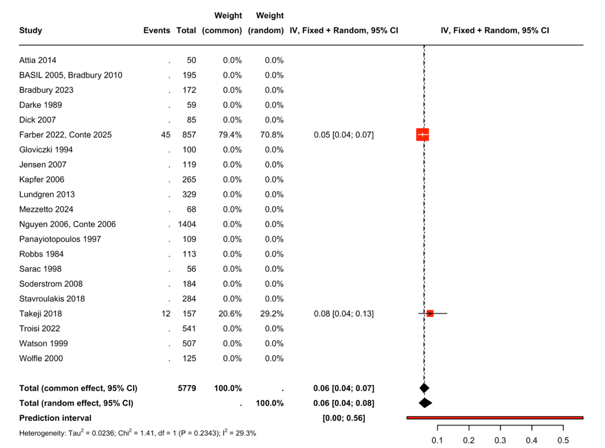

**SM2.6** Forest plot summarising the pooled prevalence of chronic kidney disease in: A) infrainguinal bypass cohorts, and B) major lower limb amputation cohorts.

**A.**

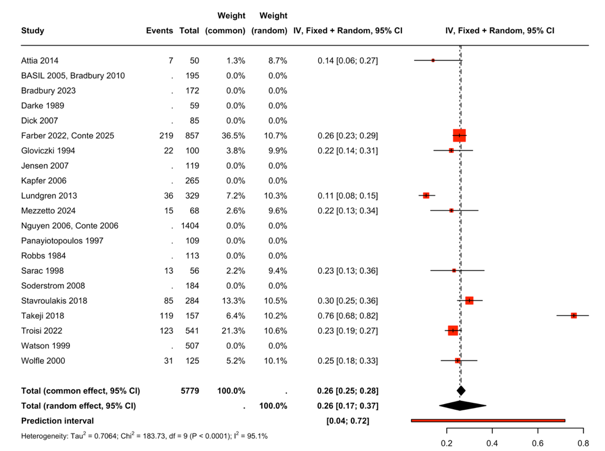

**B.**

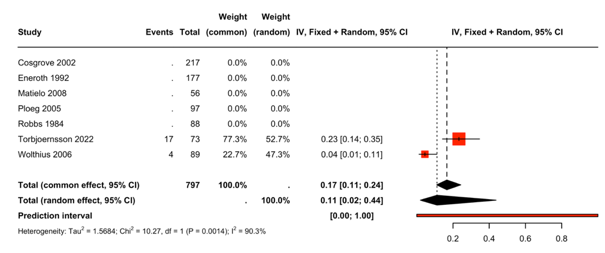

**SM2.7** Forest plot summarising the pooled prevalence of end-stage renal failure in: A) infrainguinal bypass cohorts, and B) major lower limb amputation cohorts.

**A.**

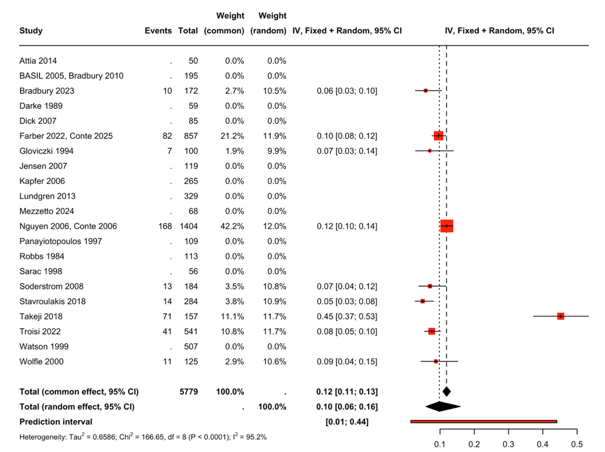

**B.**

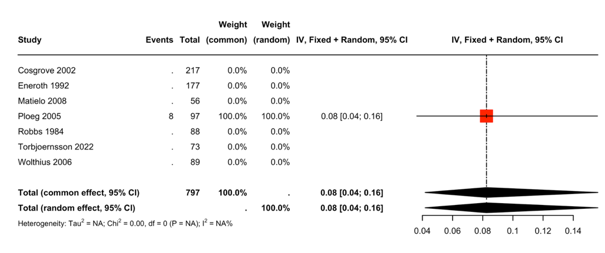

**SM2.8** Forest plot summarising the pooled prevalence of current smokers in: A) infrainguinal bypass cohorts, and B) major lower limb amputation cohorts.

**A.**

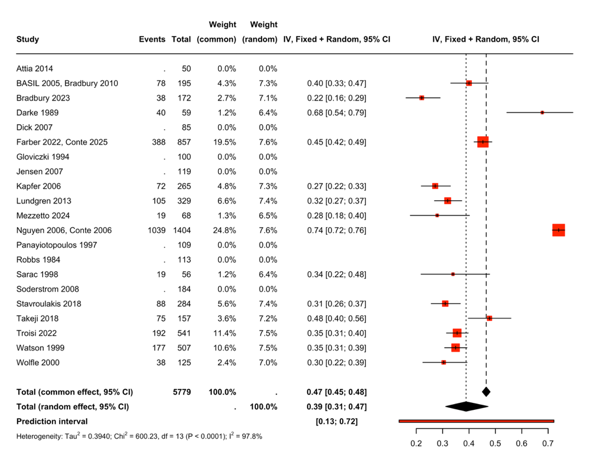

**B.**

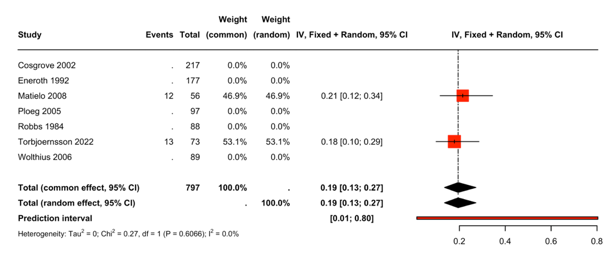

**SM2.9** Forest plot summarising the pooled prevalence of participants who have ever smoked in infrainguinal bypass cohorts. There were no studies reporting this for major lower limb amputation cohorts.

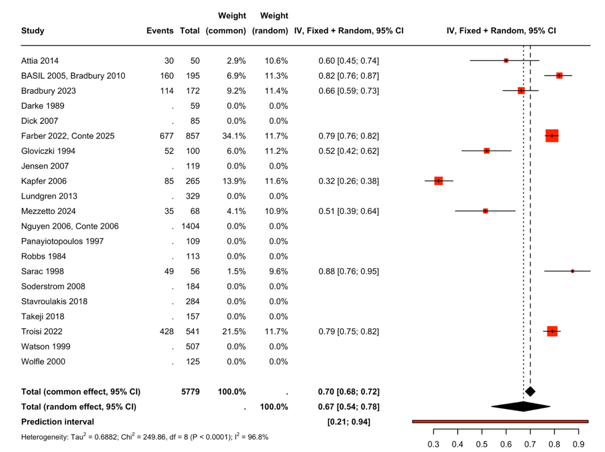

**SM2.10** Forest plot summarising the pooled prevalence of chronic obstructive pulmonary disease in: A) infrainguinal bypass cohorts, and B) major lower limb amputation cohorts.

**A.**

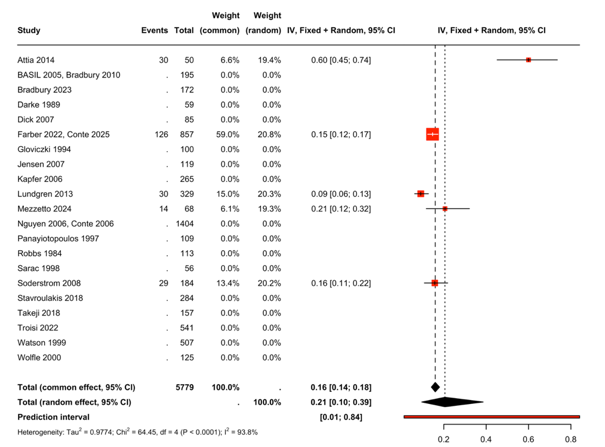

**B.**

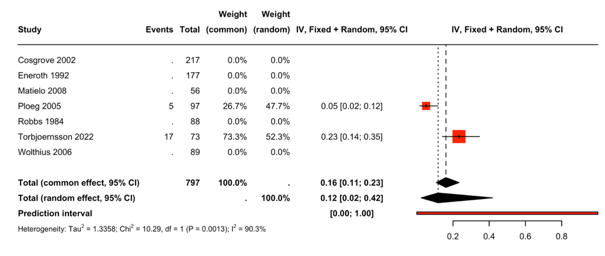
